# Genome-wide association meta-analysis of age at onset of walking

**DOI:** 10.1101/2024.05.07.24306845

**Authors:** Anna Gui, Anja Hollowell, Emilie M. Wigdor, Morgan J. Morgan, Laurie J. Hannigan, Elizabeth C. Corfield, Veronika Odintsova, Jouke-Jan Hottenga, Andrew Wong, René Pool, Harriet Cullen, Siân Wilson, Varun Warrier, Espen M. Eilertsen, Ole A. Andreassen, Christel M. Middeldorp, Beate St Pourcain, Meike Bartels, Dorret I. Boomsma, Catharina A. Hartman, Elise B. Robinson, Tomoki Arichi, David Edwards, Mark H. Johnson, Frank Dudbridge, Stephan J. Sanders, Alexandra Havdahl, Angelica Ronald

**Affiliations:** Department of Psychology, University of Essex, Wivenhoe Park, Colchester, CO4 3SQ, United Kingdom; Centre for Brain and Cognitive Development, Department of Psychological Sciences, Birkbeck University of London, London, WC1E 7HX, United Kingdom; Institute of Developmental and Regenerative Medicine, Department of Paediatrics, University of Oxford, Oxford, OX3 7TY, United Kingdom; School of Psychology, University of Surrey, Guildford, Surrey, GU2 7XH, United Kingdom; Nic Waals Institute, Lovisenberg Diaconal Hospital, Oslo, Norway; PsychGen Centre for Genetic Epidemiology and Mental Health, Norwegian Institute of Public Health, Oslo, Norway; Population Health Sciences, Bristol Medical School, University of Bristol, United Kingdom; Department of Biological Psychology, Faculty of Behavioral and Movement Sciences, Vrije Universiteit Amsterdam, van der Boechorststraat 7, 1081 BT Amsterdam, the Netherlands; Department of Psychiatry, University Medical Center of Groningen, University of Groningen, Hanzeplein 1, 9713 GZ, Groningen, the Netherlands; MRC Unit for Lifelong Health and Ageing at UCL, 1-19 Torrington Place, London WC1E 7HB, United Kingdom; Research Department of Early Life Imaging, School of Biomedical Engineering and Imaging Sciences, King’s College London, London SE1 7EH, United Kingdom; Department of Medical and Molecular Genetics, School of Basic and Medical Biosciences, King’s College London, London SE1 9RT, United Kingdom; Fetal-Neonatal Neuroimaging & Developmental Science Center, Boston Children’s Hospital, Boston, MA, United States of America; Division of Newborn Medicine, Harvard Medical School, Boston, MA, United States of America; Department of Psychiatry, University of Cambridge, Douglas House, Trumpington Road, Cambridge, CB2 8AH, United Kingdom; Department of Psychology, PROMENTA Research Center, University of Oslo, Oslo, Norway; NORMENT Centre, Institute of Clinical Medicine, University of Oslo and Division of Mental Health and Addiction, Oslo University Hospital, 0407 Oslo, Norway; KG Jebsen Centre for Neurodevelopmental disorders, University of Oslo, Oslo, Norway; Department of child and youth psychiatry and psychology, Amsterdam UMC; Amsterdam, The Netherlands; Amsterdam Reproduction and Development Research Institute, Amsterdam Public Health Research Institute, Amsterdam, the Netherlands; Arkin mental health care, Amsterdam, The Netherlands; Levvel, Academic Center for Child and Adolescent Psychiatry, Amsterdam, The Netherlands; Child Health Research Centre, University of Queensland, Brisbane, Australia; Child and Youth Mental Health Service, Children’s Health Queensland Hospital and Health Service, Brisbane, Australia; Max Planck Institute for Psycholinguistics, Wundtlaan 1, 6525 XD Nijmegen, The Netherlands; MRC Integrative Epidemiology Unit, University of Bristol, United Kingdom; Donders Institute for Brain, Cognition and Behaviour, Radboud University, The Netherlands; Department of Complex Trait Genetics, Center for Neurogenomics and Cognitive Research, Amsterdam, Vrije Universiteit, Amsterdam, The Netherlands; University Medical Center Psychopathology and Emotion regulation (ICPE), Department of Psychiatry, University Medical Center Groningen, University of Groningen, The Netherlands; Broad Institute, Boston, United States of America; Centre for the Developing Brain, School of Biomedical Engineering and Imaging Sciences, King’s College London, London SE1 7EH, United Kingdom; Department of Psychology, University of Cambridge, Cambridge, CB2 3EB, United Kingdom; Department of Population Health Sciences, University of Leicester, George Davies Centre, Leicester, LE1 7RH, United Kingdom; Department of Psychiatry and Behavioral Sciences, UCSF Weill Institute for Neurosciences, University of California, San Francisco, CA 94158, United States of America

## Abstract

Onset of walking is a developmental milestone with wide individual differences and high heritability in humans. In this genome-wide association study meta-analysis of age at onset of walking (N=70,560 European-ancestry infants), SNP-based heritability was 24.13% (SE=1.16%) with ∼11.9K variants accounting for about 90% of it, suggesting high polygenicity. We identified 11 independent genome-wide significant loci, including a “double hit” haplotype in which both decreased expression of *RBL2* and a potentially deleterious missense variant in *RBL2* are associated with delayed walking. Age at onset of walking (in months) was negatively genetically correlated with ADHD and BMI, and positively genetically correlated with intelligence, educational attainment, and adult brain gyrification. The polygenic score showed out-of-sample prediction of 3-5.6%, confirmed to be largely due to direct effects in sib-pair analyses, and was associated with volume of neonatal brain structures involved in motor control. This offers new biological insights of clinical relevance into neurodevelopment.

## Introduction

The development of bipedal ambulation is a key human characteristic. Although most humans begin to walk independently by early childhood, typical attainment of this milestone can be achieved within a relatively wide developmental period, for most infants between 8 and 18 months old^1^. It is thought that age at onset of independent walking is a complex trait determined by multiple factors, including body dimensions, year of birth, gestational age and related neural maturation, opportunity to practice^2,3^, cultural context^4^ and nutrition^5^. Many of these factors are thought to influence the structure and function of a network of brain areas implicated in motor control, including the cortex, basal ganglia, and cerebellum, with dysfunction in these brain regions resulting in movement disorders^6^.

In early childhood, the onset of walking is a simple yet robust means to measure gross motor skill development, but also indexes broader aspects of brain and behavioral development. A major advantage of this milestone is that it is both memorable and clearly defined and therefore can be reliably identified and recalled by parents^7^. Moreover, whilst there is variability in the sequence and presence of some motor skills (for example, some children bottom shuffle but never crawl), walking is an exclusive and informative milestone for both typical and atypical developmental trajectories.

In current clinical practice, an inability to walk independently by age 18 months is considered a criterion for referral for developmental delay^1^. This is because delayed walking could be due to a motor-specific issue such as a muscle disorder or generalised issues such as global developmental delay. The causes of these issues can be genetic or environmental, including genetic disorders and extreme prematurity^8^. However, historical data suggest that only a minority (about a third) of late walkers may have an underlying neurological abnormality or developmental disorder and that variation in age at onset of walking within the typical range was not strongly associated with IQ in childhood^9^. As such, late walking children (later than 18 months) might either reflect an extreme of typical variation or relate to clinically meaningful conditions with a later age of onset.

In addition to reflecting general developmental processes, the ability to walk independently may itself have cascading effects on other developmental domains^10^. The degree to which early attainment of motor milestones predicts early attainment in other domains (e.g., social or language) remains unknown. We do know that when children transition from crawling to standing and walking, the perspective at which they perceive the world changes, as do their means of interacting with the world^11^. Infants who can walk independently release their hands and consequently have more opportunities to carry and manipulate objects while moving around. However, it remains unclear what are the causal influences underlying the wide variability in age at onset of walking or whether these causal influences are also associated with later health, neurodevelopmental and cognitive outcomes.

A greater understanding of the variability and causes of late walking has clear societal implications. It would inform many countries’ public health policy that aim to screen children for delay^12^. In addition to the resulting healthcare costs for assessment and investigation, many parents will also experience undue stress if their child’s late walking leads to clinical follow up in the context of normal variation, i.e. the child’s late walking is not signalling clinical need per se, but just their predisposition for later walking. Predictive factors, such as genetic information, have the potential to offer greater understanding regarding the etiology of this developmental milestone. Furthermore, they can contribute alongside screening tools to aid the prediction and early identification of clinically-relevant conditions associated with early or delayed onset of walking, and avoid missing time for potentially beneficial physical training when appropriate.

There is substantial evidence for a genetic contribution to motor development. A recent meta-analysis of infant twin studies showed that the broad category of psychomotor function was one of the most heritable behavioural domains, with pooled heritability of 59%^13^. For age at onset of walking specifically, a study of 2,274 twin pairs in England and Wales reported a heritability of 84%^14^. Polygenic scores for autism spectrum disorder (ASD, hereafter autism), schizophrenia and bipolar disorder have been found to be associated with infant neuromotor characteristics such as muscle tone, reflexes and senses^15^. Further, the ADHD polygenic score was associated with age at onset of walking^16^. As such, age at onset of walking appears to be an ideal candidate for locus discovery research. Identification of specific genetic loci is an important step towards uncovering the biological mechanisms underlying this neurodevelopmental milestone and potentially deriving clinically-informative insights with respect to childhood motor disorders.

In sum, there are several reasons for focusing on age of walking onset. It is easily measurable in large cohorts, reliably recalled by parents^7^ and varies substantially within the typical population. It is a key milestone with potential consequences for many other aspects of physical, social and cognitive development. It is a prime example of an emergent phenomenon in human development dependent on multiple interacting preceding factors. Finally, although it is used as a clinical marker in public health, the majority of late walkers are false positives in the sense that they do not have any clinically relevant need.

Here we present the first genome-wide association study (GWAS) meta-analysis of age at onset of walking on a sample of 70,560 children from four European-ancestry cohorts. First, we found that common genetic variation explained 24% of the interindividual variability in age at onset of walking and it was a highly polygenic trait. Second, we identified 11 independent genetic loci associated with age at onset of walking, two of which colocalised with eQTLs in genes *RBL2* and *KANSL1,* respectively. Third, we found significant genetic correlations between onset of walking and physical health indicators, neurodevelopmental conditions and cognitive traits, psychiatric disorders and cortical phenotypes. Fourth, the polygenic score showed out-of-sample prediction of 3-5.6% confirmed to be largely due to direct effects in sib-pair analyses. We also found that this polygenic score was positively associated with gross motor skills at age 18 months and with the volume of neonatal brain structures involved in motor control in independent samples.

## Results

### Genomic loci associated with age at onset of walking

We conducted a GWAS meta-analysis of age at onset of walking in a sample of 70,560 children including data from four European-ancestry cohorts: Norwegian Mother, Father and Child Cohort Study^17,18^ (MoBa, N = 58,302), Netherlands Twin Register^19^ (NTR, N = 6,251), Lifelines multi-generational prospective population-based birth cohort study^20^ (N = 3,415) and Medical Research Council National Study for Health and Development^21^ (NSHD, N = 2,592). The quantile-quantile (QQ) plot for the MoBa GWAS (Supplementary Fig. S1) indicated a p-value deviation from a normal distribution (λ_GC_ = 1.23). The observed inflation is likely explained by trait polygenicity (LD Score Regression intercept = 1.008 (0.008)^22,23^, see Supplementary Note A for a detailed investigation of observed inflation). Furthermore, the other cohorts’ inflation factors were below the recommended threshold of 1.10 (NTR λ_GC_ = 0.975, Fig. S2, Lifelines λ_GC_ = 1.001, Fig. S3, NSHD λ_GC_ = 1.002, Fig. S4). Therefore, automatic correction for genomic control was not applied for all cohorts when performing the standard error-weighted meta-analysis using the METAL tool^24^.

We identified 11 independent genome-wide significant (p < 5 × 10^-8^) loci with one lead variant per locus in GCTA conditional and joint analysis (COJO)^25^ (Table 1, Fig. 1, see also Supplementary Fig. S5 for the QQ-plot and Fig. S6 for the regional plots). All 11 lead SNPs remained significant after conditioning on the other significant SNPs on the same chromosome and were the only signal within each locus. The most strongly associated SNP was located on chromosome 12 (rs7956202 near *HECTD4*, p = 2.045 × 10^-11^). This variant has been previously associated with reaction time^26^ and variants in LD with rs7956202 have been associated with physical traits such as diastolic blood pressure^27^ and hip circumference adjusted for BMI^28^ (Supplementary Table S4). The second most significant lead SNP was located on chromosome 16 (rs16952251, near *RBL2*, p = 2.637 × 10^-11^), fine mapping of this locus is discussed later (see Results section “Colocalization with gene expression in the brain”). See Table 1 for a full list of significant loci and Supplementary Table S4 for previous associations with complex traits.

**Fig. 1.**
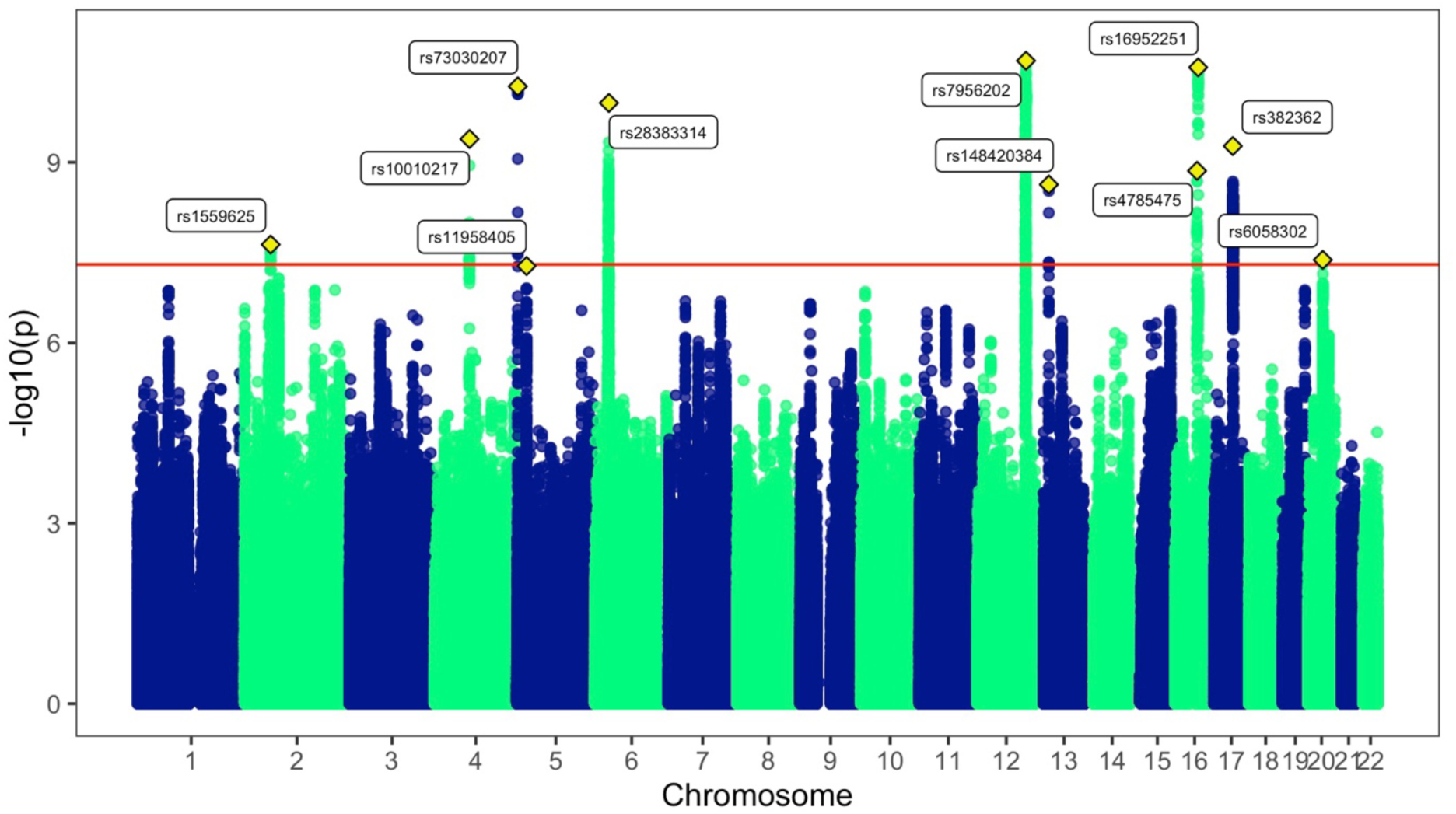
*Manhattan plot of the GWAS meta-analysis of age at onset of walking*. The x axis shows genomic position (chromosomes 1-22) and the y axis shows statistical significance as −log_10_[p-value]. P-values are two-sided and based on an inverse-variance standard-error weighted fixed-effects meta-analysis. The horizontal red line indicates the p-value threshold for genome-wide statistical significance (p = 5 x 10^-8^). The lead SNP for each genome-wide significant locus is labelled and indicated with a yellow diamond. The inflation factor λ_GC_ for this GWAS was 1.27 and LDSC intercept 1.00 (SE = 0.01), suggesting that inflation was due to polygenicity of age at onset of walking (see Supplementary Note A for a discussion). The meta-GWAS QQ-plot by allele frequency is presented on Fig. S5.

**Table 1.**
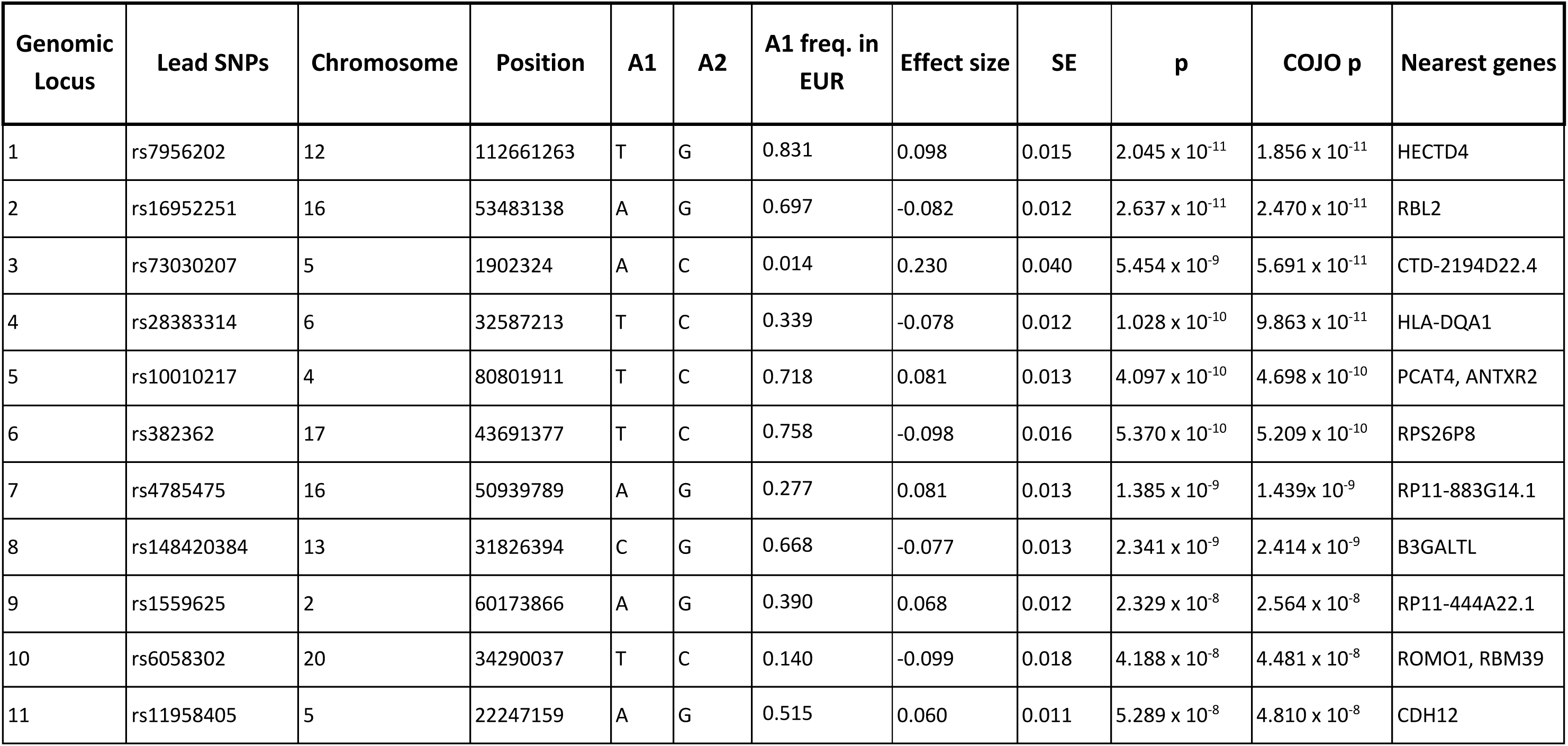
Genome-wide significant loci associated with age at onset of walking. The allele frequency in the 1000 Genomes^29^ European-ancestry sample (EUR), the effect sizes and the standard errors (SE) refer to Allele 1 (A1). The p-values of association from the meta-analysis performed in METAL and p-values resulting from the conditional and joint (COJO)^25^ analysis are reported. The nearest genes were identified using FUMA^30^.

### Common genetic architecture of age at onset of walking

SNP-based heritability of age at onset of walking estimated with LDSC^22^ was h^2^_SNP_ = 24.13% (SE = 1.16%, 95% CIs [21.86 - 26.40]). Heritability for the phenotype in males and females h^2^_SNP_ were estimated to be 23.06% (SE = 1.81%, 95% CIs [19.51 - 26.61) and 23.06% (SE = 1.89%, 95% CIs [19.36 - 26.76]), respectively. The genetic correlation (r_g_) of the phenotype between males and females was 0.99 (SE = 0.06).

There was no statistically genome-wide significant heterogeneity between cohorts as tested with I^2^ (maximum I^2^ = 95.3 for SNPs rs7864115 and rs148684045, χ^2^(1) = 21.453 and 21.441, p = 3.63 x 10^-6^ and 3.65 x 10^-6^, respectively), indicating that variation of effects between individual GWASs was due to chance rather than heterogeneity between the cohorts^31^ (Fig. S7, see Supplementary Note C for genetic correlation between cohorts).

### Biological annotation of associated loci and genes

#### Analyses on prioritised genes annotated to significant SNPs

The genome-wide significant SNPs were mapped to 233 genes based on genomic position, expression quantitative trait loci (eQTLs) and chromatin interaction information in FUMA^30^ (Table S5). We tested these prioritised genes were differentially expressed in the brain across BrainSpan^32^ developmental stages and GTEx v8^33^ tissues. We observed a significant down-regulation of the differentially expressed genes (DEGs) in multiple tissues including the brain (amygdala and hippocampus) and the heart left ventricle, and DEGs up-regulation in fibroblasts (Fig. S8). The enrichment of up-regulated or down-regulated DEGs across BrainSpan developmental stages was not significant. Gene sets associated with age at onset of walking were enriched in the Gene Ontology^34^ neurogenesis and generation of neurons pathways (see Table S6 for all the significantly enriched gene sets and gene sets-trait associations from previous studies).

#### Genes associated with age at onset of walking

The MAGMA^35^ gene-based test performed in FUMA on the meta-GWAS summary statistics indicated 50 genes which were associated with age at onset of walking at a Bonferroni corrected genome-wide significance threshold of 2.664 × 10^-6^ (p = 0.05 / 18,766, Table S7). A full list of previously reported genome-wide associations with complex traits for the 50 WALK-associated genes is provided in Table S8.

Using Genomics England PanelApp^36^, we found that 13 of the 47 genes with Ensembl IDs (27.6%) were associated with intellectual disability (ID, v5.557); this is over double the proportion (2.16 times) of ID-associated genes in the panels as a whole (2,624 out of 19,950, 12.9%; p = 0.005, Chi-squared). These genes include *ATXN2*, *AUTS2*, *CUX2*, *FOXP1*, *KANSL1*, and *RBL2* (Table S7). Furthermore, we found 7 of the 47 genes were associated with autism (14.9%), which is over four times the proportion of autism-associated genes in the panel (v0.36, largely based on SFARI-gene) as a whole (734 out of 19,950, 3.67%; p = 0.0004, Chi-squared).

To identify tissue specificity of age at onset of walking, MAGMA^35^ gene-property analyses performed in FUMA using gene-based association p-values for all the 18,766 genes revealed that gene expression was primarily enriched in the brain cerebellar hemispheres and cerebellum, although this result was not significant (see Fig. S9). Overall, expression of the genes associated with age at onset of walking were significantly enriched between 19 and 24 (late mid-prenatal period) post-conceptional weeks (Fig. S10). The MAGMA gene-set analysis yielded no significant results (Table S9).

#### Analyses on the meta-GWAS summary statistics

Enrichment of age at onset of walking meta-GWAS signal by functional genomic annotation was tested using stratified LDSC^37^ analyses. These revealed that heritability of age at onset of walking was significantly enriched in genomic regions conserved in primates (17.98-fold enrichment, p = 4.68 × 10^-7^), mammals (14.58-fold enrichment, p = 2.90 × 10^-6^) and vertebrates (9.76-fold enrichment, p = 9.17 × 10^-6^, see Fig. 2A). Full results of partitioned heritability by functional genomic annotation can be found in Table S10.

**Fig. 2.**
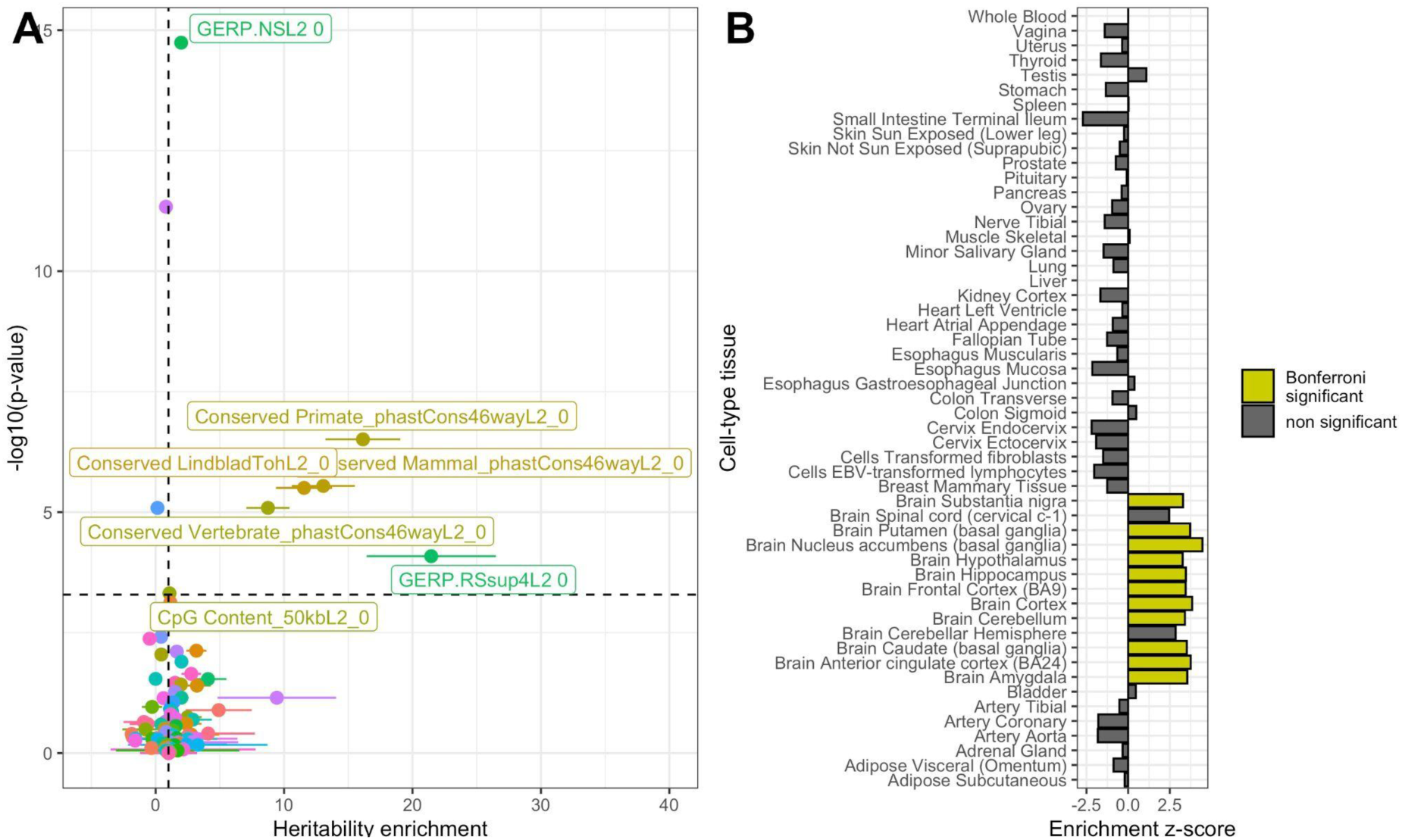
*Partitioned heritability enrichment by functional annotation and cell type* **A.** Enrichment of age at onset of walking GWAS signal by functional genomic annotation. Points represent the heritability enrichment estimate and error bars enrichment standard errors. The dashed horizontal line represents statistical significance based on Bonferroni correction for multiple testing (Supplementary Table S10). Genomic annotations with significant enrichment for age at onset of walking are labelled. Dots are colored using a spectrum of colors based on alphabetical order. **B.** Tissue enrichment based on LDSC partitioned heritability. Statistically-significant enrichments are highlighted as yellow bars.

We then tested whether heritability was enriched in specific cell types using stratified LDSC^38^ and found significant enrichment in the brain, particularly in the basal ganglia, cortex, amygdala and cerebellum (Fig. 2B). Complete stratified LDSC by cell type estimates are reported in Table S11.

### Colocalization with gene expression in the brain

We investigated whether the 11 genome-wide significant loci (p < 5 x 10^-8^)^39^, as well as 50 genes significantly associated with age at onset of walking (Table S7), were enriched for expression quantitative trait loci (eQTLs) for nearby genes in an independent dataset of post-mortem bulk RNA-seq from 261 samples of the human adult cerebellum^39^. We identified significant eQTLs for the gene *RBL2* (which encodes a transcriptional regulator by the same name) in genomic locus 2 on chromosome 16 (Table 1). Comparing the statistical evidence of association with age at onset of walking (GWAS) against the statistical evidence of association with *RBL2* expression, we noticed a distinct pattern; both the GWAS and eQTL p-values had two groups of significantly associated SNPs distinguished by their linkage disequilibrium correlation with a lead GWAS SNP (rs17800727, Fig. 3A). Group 1 had the strongest evidence for GWAS association (min p = 2.95 x 10^-11^) but slightly weaker evidence of eQTL association (min p = 2.72 x 10^-13^ cerebellum eQTL), while Group 2 had weaker evidence for GWAS association (min p = 9.51 x 10^-8^) but stronger evidence of eQTL association (min p *=* 6.41 x 10^-24^ cerebellum eQTL, Fig. 3A). We investigated the probability that the same SNPs in this locus influence both age at onset of walking and *RBL2* expression (colocalization, Fig. 3). Our colocalization analysis at this locus suggested an independent causal variant in the GWAS (rs17800727; chr16:53481010:A:G GRCh37; chr16:53447098:A:G GRCh38) and the eQTL data (rs7203132; chr16:53429775:G:A GRCh37; chr16:53395863:G:A GRCh38) with a posterior probability of 0.96^40^ that the causal SNP is distinct in each dataset.

**Fig. 3.**
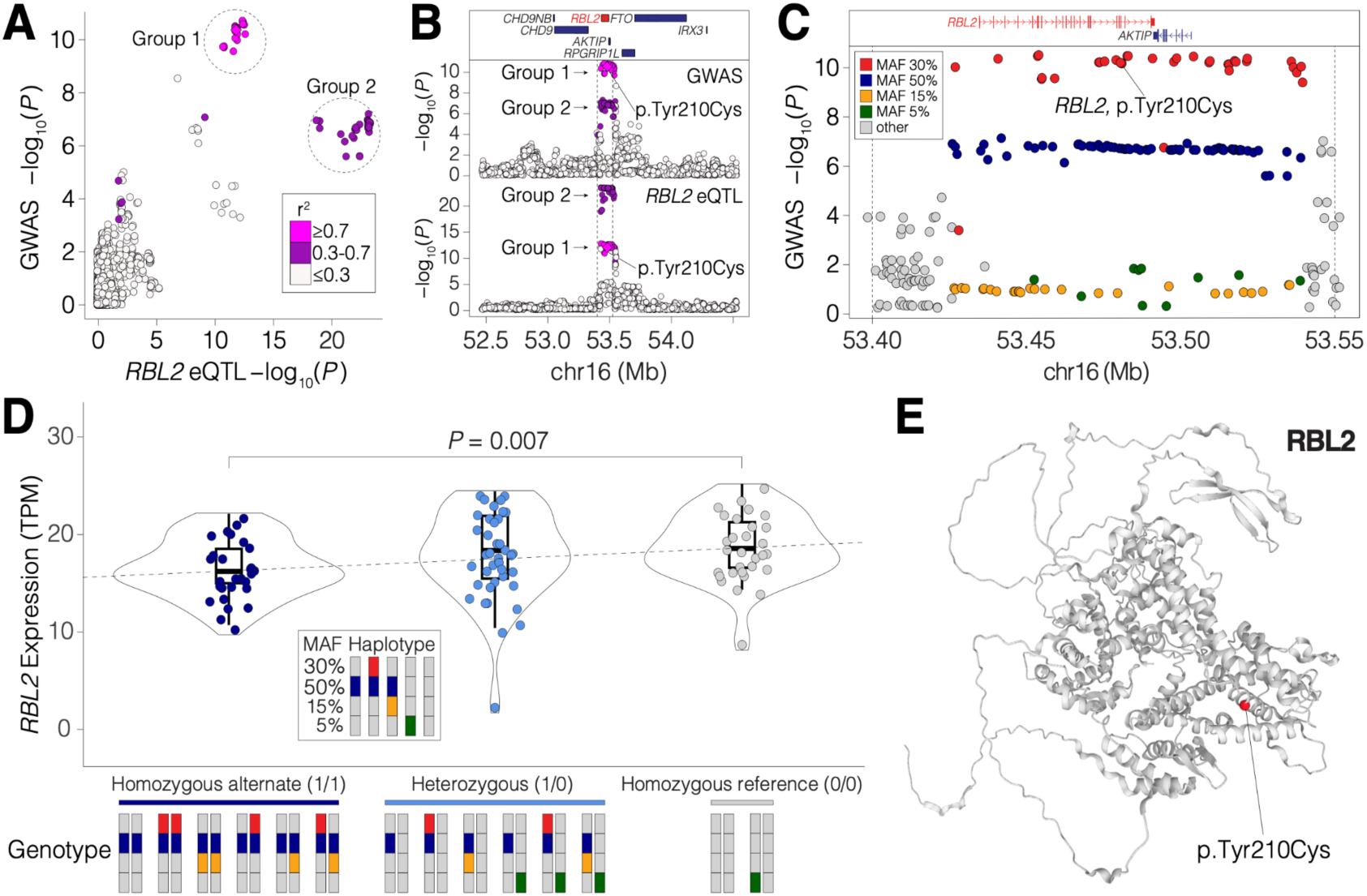
*Colocalization of variants in genomic locus 2*. Genomic locus 2 overlaps with a region in which SNPs are predicted to alter *RBL2* expression in the human brain (eQTLs). **A.** The GWAS evidence for association with age at onset of walking (-log_10_(*P*), *x*-axis) is plotted against the statistical evidence of being an eQTL for *RBL2* in human adult cerebellum^1^ (-log_10_(*P*), *y*-axis) for each SNP (points) within a 2Mb window around the GWAS peak. Points are colored by linkage disequilibrium (LD) correlation with the lead SNP (rs17800727) and these values are used to define two groups. **B.** The SNPs from ‘A’ are shown in the 2Mbp genomic region (*x*-axis, GRCh37) with protein-coding genes (top), GWAS evidence for association with age at onset (-log_10_(*P*), middle) and statistical evidence for *RBL2* expression in human cerebellum^39^ (-log_10_(*P*), *y*-axis, bottom). Point color matches ‘A’. **C.** A zoomed in view of the peak indicated by dashed vertical lines in ‘B’ shows the GWAS evidence for association with age at onset of walking (-log_10_(*P*), *y*-axis) by genomic position (*x*-axis, GRCh37). Color indicates the Minor Allele Frequency (MAF) of each SNP. The locations of protein-coding genes in the region are indicated at the top. A SNP (rs17800727) that results in a missense variant (p.Tyr210Cys) in *RBL2* is marked. **D.** Swarm, violin, and box-plots showing the distribution of *RBL2* expression (transcripts per million (TPM), *y*-axis). Each point represents the expression of *RBL2* in one of 87 prenatal human cortices (BrainVar^41^) split by genotype into three groups based on zygosity for the Group 2 50% MAF SNPs. The *P*-value represents the difference between the homozygous alternate and homozygous reference groups (Wilcoxon, two-sided). Bars at the bottom indicate pairs of haplotypes (derived from the data shown in ‘C.’ making up each genotype. **E.** Structure of RBL2 protein predicted by AlphaFold^45^ with the location of rs17800727, p.Tyr210Cys in red^46^.

To understand these two groups, we assessed their distribution across the 2Mb genomic locus (± 1 MB around the gene) and observed that they overlapped throughout a 125kb peak with well-defined margins for both the GWAS and *RBL2* eQTL analysis (Fig. 3B). We next considered how these SNPs were distributed based on minor allele frequency (MAF, Fig. 3C). The Group 1 SNPs (strongest GWAS evidence) had a MAF of 30%, while the Group 2 SNPs (strongest eQTL evidence) had a MAF of 50%. Using whole-genome sequencing data from 176 individuals with paired *post-mortem* RNA-seq data from prefrontal cortex^41^, we used the MAF distribution to identify five haplotypes (Fig. 3D) and each individual’s genotype. Group 2 SNPs (strongest eQTL evidence, MAF 50%) are found in three haplotypes (dark blue and red, dark blue and yellow, dark blue alone, Fig. 3D) resulting in the high MAF of 50%. Homozygous status for the Group 2 SNPs is associated with decreased expression of *RBL2* (p = 0.007, Wilcoxon). We infer that one of the SNPs shown in dark blue (Fig. 3C) impacts *RBL2* expression, though no clear candidate SNP was evident when considering epigenetic data.

Group 1 SNPs are only found on one haplotype (dark blue and red, Fig. 3D) resulting in a lower MAF of 30% than the Group 2 SNPs. We infer that one of the Group 1 SNPs has a functional impact above and beyond the decrease in *RBL2* expression mediated by the Group 2 SNPs, to yield the stronger evidence of association with age at onset of walking. Annotation of the 125kb locus with VEP^42^ identified rs17800727 as a likely candidate for this effect, since it results in a missense variant (MANE isoform: ENST00000262133.11, p.Tyr210Cys) (Fig. 3E) that is predicted to impact function by some severity metrics (e.g., ‘Damaging’ based on PolyPhen2^43^, CADD^44^ score of 25) but not all (e.g., ‘Tolerated’ based on SIFT); future functional studies would be required to validate this functional impact.

The presence of the rs17800727 *RBL2* missense variant on a haplotype associated with decreased *RBL2* expression (red co-occurs with dark blue, Fig. 3D) would be a parsimonious explanation for the two observed groups of SNPs. Group 2 SNPs reflect three haplotypes that decrease *RBL2* expression, resulting in stronger eQTL association than association with age at onset of walking (dark blue, dark blue and red, dark blue and yellow, Fig. 3D). Group 1 SNPs reflect one of these three haplotypes (dark blue and red, Fig. 3D). An *RBL2* missense variant in the Group 1 SNPs (red, Fig. 3C) leads to the stronger evidence in the GWAS for association with age at onset of walking (Fig. 3B). If the missense variant had a loss-of-function effect it would be on a haplotype that magnifies the functional impact through decreased expression of *RBL2*.

We also identified colocalization of SNPs associated with expression of *KANSL1* in the cerebellum with SNPs associated with age at onset of walking in genomic locus 6 on chromosome 17 (Table 1). Colocalization analysis^40^ identified rs1078268 as the shared causal variant in both GWAS and eQTL datasets at this locus (PP=0.79; chr17:44075901:A:G GRCh37; chr17-44075901-A-G GRCh38; Fig. S11). The alternate allele (G) for this variant is associated with increased age at onset of walking and increased expression of *KANSL1*.

### Polygenic score analysis

In a leave-one-out design, we calculated a polygenic score (PGS) based on meta-analyses of all samples, leaving out either Lifelines, NTR or NSHD. In the Lifelines cohort, the PGS from the meta-GWAS of the other cohorts (MoBa, NTR and NSHD) was significantly associated with age at onset of walking (β = 0.19, SE = 0.02, p < 2 × 10^-16^, R^2^ = 0.034). Using the same method, the PGS was significantly associated with age at onset of walking in the NTR cohort (β = 0.18, SE = 0.02, p < 2 × 10^-16^, R^2^ = 0.032) and in the NSHD cohort (β = 0.18, SE = 0.02, p < 2 × 10^-16^, R^2^ = 0.030). The MoBa sample comprised a high proportion of the data such that it would be inappropriate as a “left out” sample in a leave-one-out design. Therefore, we applied five-fold cross-validation to this cohort, yielding five within-sample PGS with a mean variance explained of R^2^ = 0.056 (SE = 0.001).

Genetic effects identified by GWAS can be confounded by indirect genetic effects, for example through population structure, assortative mating and passive gene-environment correlation (prGE)^47^. To identify possible confounding from indirect genetic effects, we used a within- and between-sib-pair PGS analysis. We generated a PGS from a meta-analysis of the MoBa, Lifelines and NSHD GWAS summary statistics and used to conduct within-family associations in the NTR dataset. Among 2,586 dizygotic twin pairs, within- and between-family standardised regression coefficients in a linear mixed-effects model were not significantly different from each other (χ^2^(1) = 0.04, p = 0.70), indicating that the genetic signal is not biassed by prGE, or effects such as stratification and assortative mating. Fig. 4 shows the beta estimates of the age at onset of walking PGS prediction in all the cohorts.

**Fig. 4.**
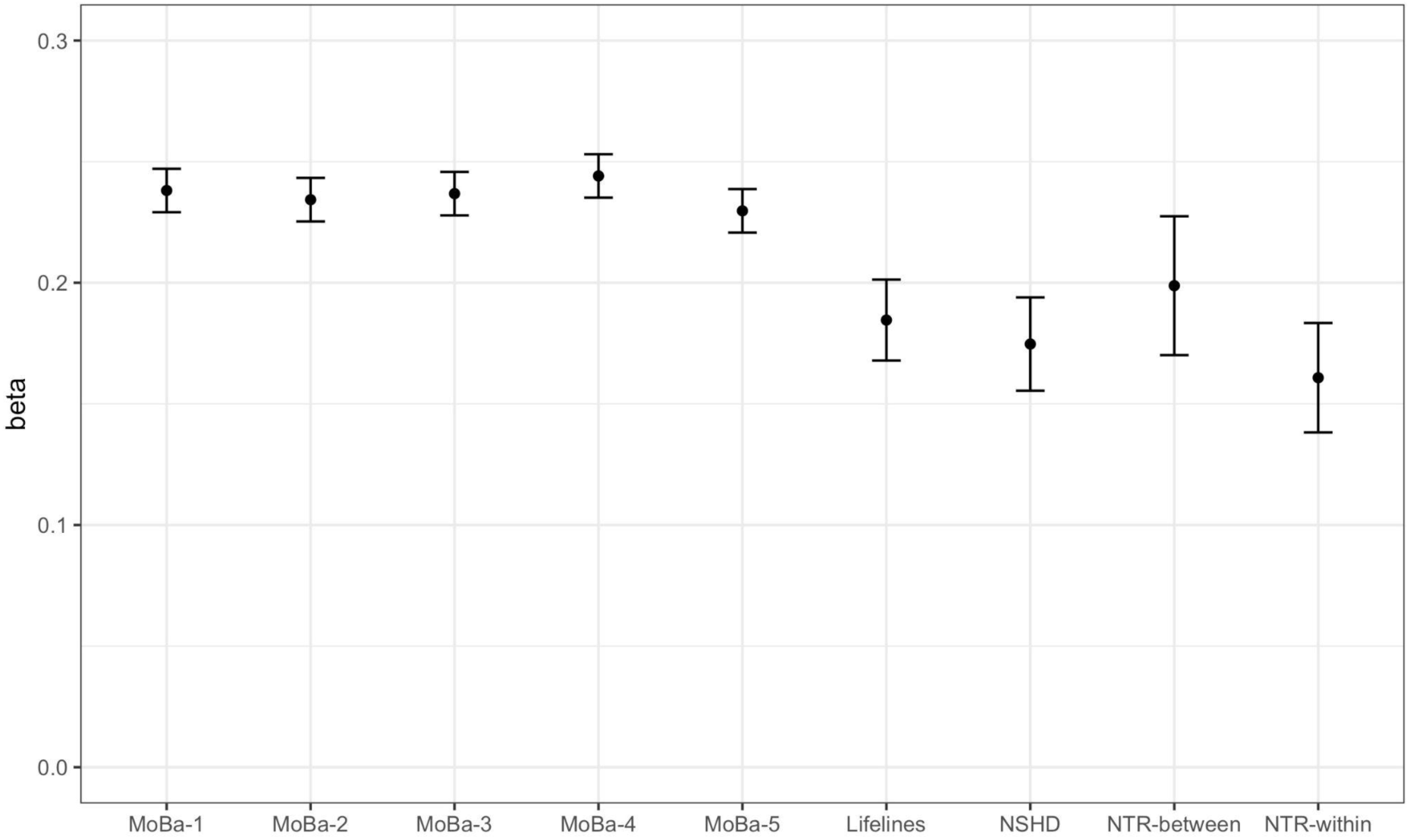
*Beta estimates of the prediction of age at onset of walking for the five MoBa subsamples, Lifelines, NSHD, NTR between and NTR within sib-pair polygenic score analysis.* Error bars represent the standard error of the beta estimate.

### Genetic correlations with other traits

Next, we tested for genetic correlations between age at onset of walking and a pre-registered selection of physical health, neurodevelopmental, psychiatric, cognitive and cortical phenotypes. Age at onset of walking was negatively genetically correlated with childhood BMI^48^ (r_g_ =-0.14, SE = 0.04, 95%CI [-0.22, -0.07]), adult BMI^49^ (r_g_ = -0.10, SE = 0.02, 95%CI [-0.14, -0.06]) and ADHD^50^ (r_g_ = -0.18, SE = 0.03, 95%CI [-0.24, -0.12]) and positively genetically correlated with educational attainment^51^ (r_g_ = 0.12, SE = 0.02, 95%CI [0.08, 0.16]), IQ^52^ (r_g_ = 0.09, SE = 0.03, 95%CI [0.04, 0.14]), and bipolar disorder^53^ (r_g_ = 0.07, SE = 0.02, 95%CI [0.03, 0.12]).

Among thirteen adolescent and adult cortical phenotypes^54^, we observed a significant genetic correlation between age at onset of walking and folding index (r_g_ = 0.14, SE = 0.04, 95%CI [0.06, 0.21]), the local gyrification index (r_g_ = 0.10, SE = 0.04, 95%CI [0.03, 0.17]) and cortical surface area (r_g_ = 0.07, SE = 0.04, 95%CI [0.002, 0.15]), all of which are measures of cortical expansion, and isotropic volume fraction, that indicates water diffusion in the brain related to ventricles and cerebrospinal fluid (r_g_ = 0.10, SE = 0.04, 95%CI [0.01, 0.17]). There were no significant genetic correlations with the other complex traits tested (see Supplementary Table S12 and Fig. 5A). Non pre-registered exploratory analyses showed that age at onset of walking was genetically correlated with self-reported walking pace in adults (r_g_ = 0.06, SE = 0.03, 95%CI [0.01, 0.11]), but not with other motor phenotypes (Table S12).

**Fig. 5.**
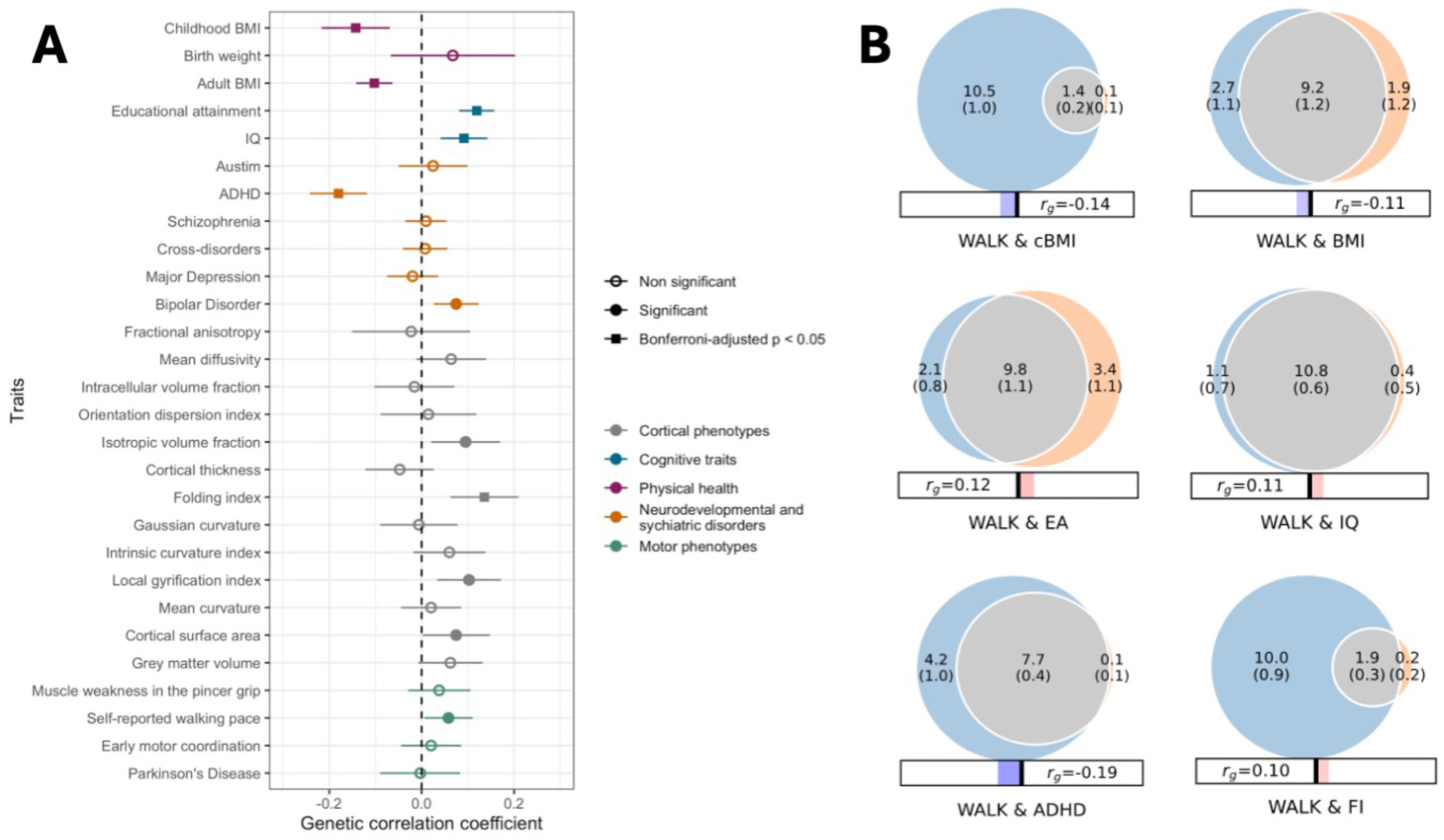
*Genetic overlap between age at onset of walking and other complex traits*. **A** Genetic correlation between age at onset of walking and physical health (purple), neurodevelopmental conditions and cognitive traits (blue), psychiatric disorders (orange), cortical (grey) and non pre-registered motor phenotypes (green). Error bars indicate 95% confidence intervals (CIs). Significant correlations based on CIs are marked with the full point. Correlations that remain significant after adjusting for multiple testing using the Bonferroni correction are marked with a squared full point. **B** Venn diagrams representing MiXeR bivariate analyses between age at onset of walking and the 9 other phenotypes with which it has significant genetic correlations. The size of the circles and the numbers within them represent the relative polygenicity of each trait (i.e., how many genetic variants contribute to 90% of the SNP heritability). The overlap between each pair of circles represents the degree of genetic overlap between the two phenotypes, that is, the number of shared variants in thousands, along with the standard error. Numbers and standard errors in sections of the circles that do not overlap represent the number of variants unique to that phenotype. The corresponding r_g_, estimated using LDSC, is shown below each Venn diagram. Bivariate results are shown for age at onset of walking (WALK), with Educational Attainment (EA), adult Body-Mass Index (BMI), Attention Deficit/Hyperactivity Disorder (ADHD), Intelligence (IQ), childhood Body-Mass Index (cBMI) and brain Folding Index (FI).

In a genetic multivariable regression performed with GenomicSEM^55^, we observed that the relationship between the genetic components of ADHD and age at onset of walking remained significant after conditioning for educational attainment (standardized β = -0.160, SE = 0.045, p = 3.8 x 10^-4^), while the conditional standardized association between educational attainment and age at onset of walking was non-significant (β = 0.038, SE = 0.033, p = 0.246, Fig. S12).

We applied MiXeR univariate and bivariate Gaussian mixture modelling^56^, which calculates the polygenicity of age at onset of walking, as the number of SNPs that explain 90% of the h^2^_SNP_, and the genetic overlap between age at onset of walking and other phenotypes, including SNPs of both concordant and discordant effect directions. We applied bivariate mixture modelling to age at onset of walking with all other phenotypes with which there was a significant genetic correlation as calculated by LDSC, after correction for multiple testing (as per Fig. 5A). MiXeR bivariate models were only applied if the phenotype met the criterion of h^2^_SNP_ x GWAS sample size > 6,000^57^. The BIC values did not support the bivariate models between age at onset of walking and local gyrification index, isotropic volume fraction and bipolar disorder; for the bivariate models with childhood BMI and folding index, BIC values did not support the models but the AIC values did, so these results should be interpreted with caution. For all other models, we found support (as evaluated using BIC), for the MiXeR model above the ‘minimal model’, which contains the minimum polygenic overlap needed to explain the LDSC genetic correlations. AIC and BIC values for all models, and negative log-likelihood plots are provided in Supplementary Table S13 and Supplementary Fig. S13 respectively.

The polygenicity of age at onset of walking was 11,857 SNPs, confirming the hypothesis that the inflation observed in the QQ-plot could be explained by trait polygenicity (Supplementary Note A). MiXeR presents the genetic overlap between two traits as Venn diagrams (Fig. 5B). In terms of the proportion of the SNPs contributing to the polygenicity of age at onset of walking that overlap with other phenotypes investigated, the traits investigated that showed the most overlap were educational attainment and IQ (82.44% and 91.07% respectively). Of these overlapping SNPs between age at onset of walking and educational attainment and IQ, the fractions of SNPs that had concordant directions of effect were 55.10% and 53.70% for educational attainment and IQ respectively. A summary of all the bivariate MiXeR analysis results can be found in Table 2.

**Table 2.** MiXeR results showing the composition of the genetic overlap between age at onset of walking and 9 traits for which there was a significant genetic correlation as measured using LDSC. Fraction shared = the proportion of the SNPs that explain 90% of the age at onset of walking h^2^_SNP_ that are shared with the correlated trait. Fraction concordant within shared = the proportion of the shared SNPs that have concordant effect direction between the two traits. r_g_ = LDSC genetic correlation as calculated within the MiXeR bivariate analysis software. The r_g_ values in Table 2 (LDSC r_g_ values reported by MiXeR) can vary slightly from those reported above because of subtle differences in the pre-processing of summary statistics prior to analysis. The magnitude of the estimates of the fraction of shared SNPs from MiXeR bivariate analyses do not correlate positively with the magnitude of the genetic correlation estimated by LDSC because the fraction of shared SNPS from MiXeR includes SNPs that have both concordant and discordant effect directions between the two traits, which counteract each other in the calculation of r_g_ by LDSC.

| Correlated trait | Fraction shared (%) | $r_g$ (M) | $r_g$ (SD) | Fraction concordant within shared (M) | Fraction concordant within shared (SD) |
| --- | --- | --- | --- | --- | --- |
| Educational attainment | 82.44% | 0.124 | 0.007 | 55.10% | 0.57% |
| IQ | 91.07% | 0.109 | 0.008 | 53.71% | 0.26% |
| Adult BMI | 77.38% | -0.112 | 0.010 | 45.46% | 0.73% |
| ADHD | 64.87% | -0.191 | 0.009 | 42.32% | 0.51% |
| Folding index | 15.84% | 0.100 | 0.016 | 58.72% | 2.12% |
| Childhood BMI | 11.80% | -0.137 | 0.015 | 36.44% | 2.38% |

### Polygenic score association with measurable differences in brain volume at birth

In an exploratory analysis we tested whether the PGS for age at onset of walking was associated with measurable differences in infant brain volume at birth. We used neonatal T2 imaging data from a European subsample of 264 term-born infants (137 male, 127 female), acquired as part of the Developing Human Connectome Project (dHCP)^58^.

The effect of the age at onset of walking PGS on brain volume was investigated across the whole-brain at the voxel-level using log-Jacobian determinants, calculated using non-linear deformation fields between subjects and the dHCP neonatal standardised atlas. In the resultant maps, higher log-Jacobian values represent brain regions that contracted during image registration (i.e., larger brain volumes), while smaller values represent volume reductions^59^. We performed a tensor-based morphometry (TBM) analysis, applying a general linear model (GLM) and permutation testing for statistical inference. We found a significant positive correlation between the age at onset of walking PGS and regional brain volume in the right basal ganglia, right posterior thalamus, bilateral anterior thalami, bilateral cerebellum and cerebellar peduncles, pons, medulla, primary visual cortex and superior temporal sulcus after correcting for multiple comparisons and thresholding at a corrected p < 0.05 (Fig. 6). Increased brain volume in these regions was associated with a higher PGS (predisposing for later age at onset of walking).

**Fig. 6.**
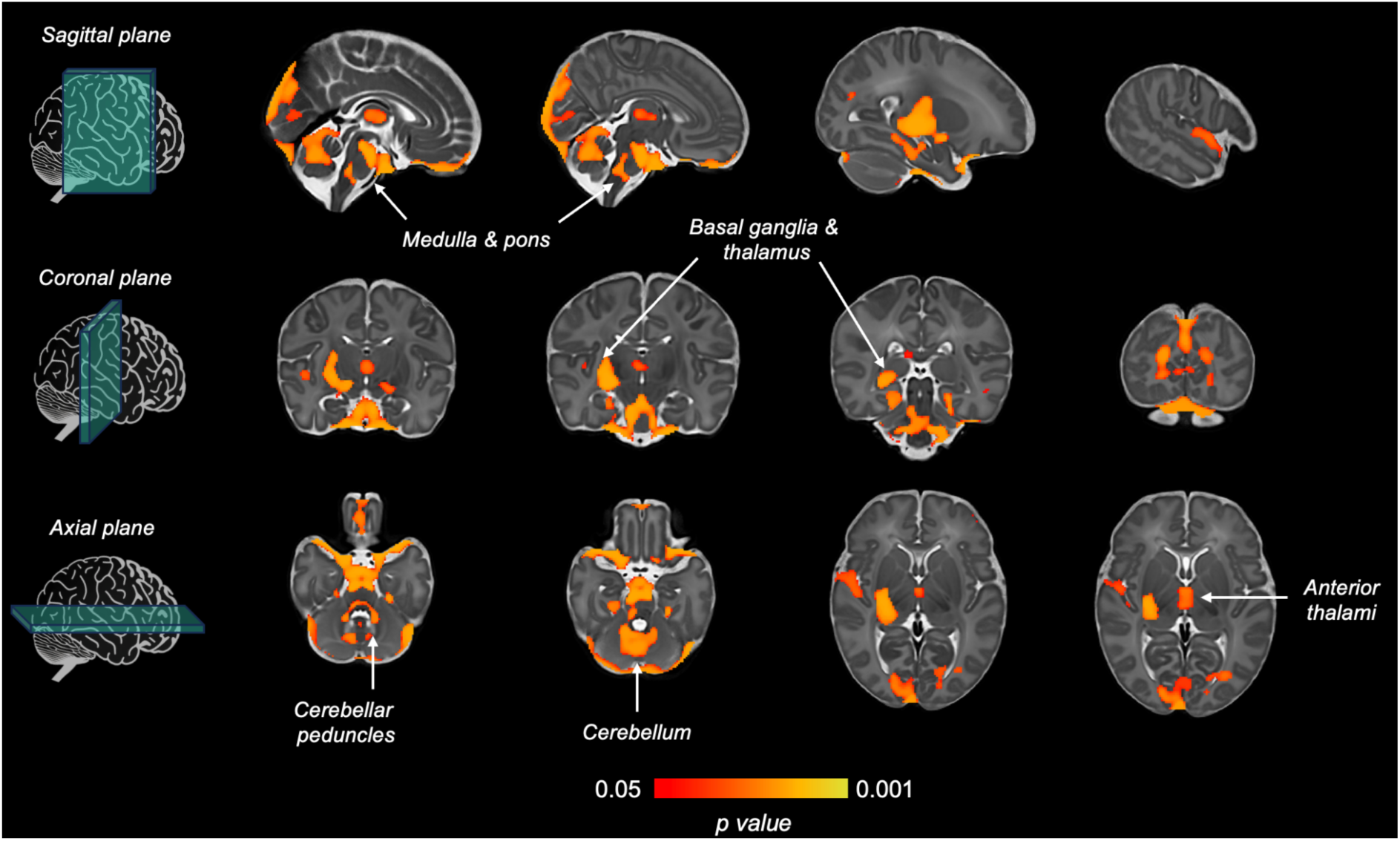
Brain regions where there is a statistically significant positive correlation between tissue volume and age at onset of walking polygenic score. Thresholding t-statistic image at t > 0.95, significant voxels were overlaid on the 40-week neonatal brain template in sagittal, coronal and axial planes. White arrows indicate significant brain structures involved in motor control.

Finally, for the European term-born infants in dHCP infants that had also been assessed using the Bayley-III Scales of Infant and Toddler Development^60^ at an age 18-month follow-up (N = 217), we explored the relationship between scaled gross motor score and the age at onset of walking PGS using a regression model. Sex, gestational age at birth, birth weight z-score, home environment score (as a proxy for socioeconomic status) and 10 ancestral PCs were included as covariates. We found that higher age at onset of walking PGS was significantly associated with lower Bayley’s gross motor score, indicating worse/possibly delayed gross motor skills (β = -0.161, SE = 0.070, p = 0.022).

## Discussion

The present study identified age at onset of walking as a polygenic phenotype with significant SNP-based heritability of 24%. We identified 11 independent genome-wide significant loci in this first GWAS of age at onset of walking, based on a sample size of 70,560 children, and these included 2 loci that colocalised with eQTLs. We discuss four main conclusions from these results.

Past models of gross motor skills, as well as neurodevelopment more generally, have put a primary emphasis on environmental factors such as nutrition^5^ and cultural factors^4,61^. Our results show for the first time that the phenotype of age at onset of walking is also associated with common genetic variants operating in the brain.

Significantly enriched cell type tissues were exclusively brain-based tissues, and moreover, strongest signals included tissues in the basal ganglia, cortex and cerebellum. In line with these findings, the polygenic score for age at onset of walking was associated with neonatal brain volume of the basal ganglia, thalami, medulla, pons and cerebellum. This is consistent with the known role of these brain areas in motor function^6,62^. Moreover, no non-brain tissue types showed significant enrichment. Based on our results, if other cell types are associated with age at onset of walking (e.g., adipose subcutaneous tissue or musculoskeletal), their involvement does not appear to be influenced by common genetic variation.

Second, the novel loci that were discovered here involve genes of highly plausible biological relevance to the onset of walking. We identify common variant association with age at onset of walking at a locus overlying Retinoblastoma-like protein 2 (*RBL2*, ENSG00000103479, genomic locus 2 in Table 1). Assessing the allele frequencies in a population, we identify five haplotypes, three of which are associated with decreased *RBL2* expression in the human cortex and one of these three also contains an *RBL2* missense variant (rs17800727). This haplotype therefore represents a “double hit”, both decreasing *RBL2* expression and including an *RBL2* missense variant. This locus has also been associated with intelligence^63^, educational attainment^64^, and height^49^; assessing the top SNP in each of these GWAS, we find they are on the same *RBL2* eQTL/missense haplotype. Based on gnomAD ^65^ v4.0, this haplotype is observed at 24-30% allele frequency in cohorts from Europe and the Middle East, 10% in South Asia, and <5% in East Asia and Africa or African Americans ^65^.

*RBL2* is also associated with an autosomal recessive neurodevelopmental disorder (eponym Brunet-Wagner)^66,67^. Homozygous loss of *RBL2* has been observed in five individuals across three families, each with a different allele ^66,67^. Affected individuals had infantile hypotonia, severe developmental delay, delayed/absent walking, and were minimally verbal. Seizures were reported in three cases. Three cases had microcephaly (-2.4SD to -4.7SD), while two had normal head circumference (65th and 50th centiles) but cerebral atrophy on MRI. Height was normal for two cases, unreported for one, and low for two (3rd centile, -3.4SD). In Balb/c mice, homozygous loss of *Rbl2* is embryonic lethal with a disorganized neural tube and neuronal loss^67^.

Heterozygous loss of *RBL2* has not been associated with a human phenotype and is observed in 0.03% of the population in gnomAD^16^. This is about half the expected number of *RBL2* null alleles, suggesting modest selective constraint, as has been observed for genes associated with other recessive disorders (LOEUF = 0.60, gnomAD^65^ v4.0).

Through colocalization analysis^40^ we also found evidence to support a shared causal variant (rs1078268) affecting both age at onset of walking and expression of *KANSL1*. *KANSL1* encodes a protein of the same name which is a conserved regulator of the chromatin modifier KAT8^68^. Heterozygous deletions at chromosome 17q21.31, that include the *KANSL1* gene, lead to an autosomal dominant neurodevelopmental disorder (eponym Koolen-de Vries), characterised by psychomotor delay in all patients, with delayed age at independent walking, as well as hypotonia, epilepsy, developmental delay/intellectual disability, congenital malformations in multiple organ systems and a characteristic facial dysmorphism^69^. Haploinsufficiency of *KANSL1* is sufficient to phenocopy the 17q21.31 microdeletion syndrome^68^. Thus, our results bring together, via genetic loci with different inheritance patterns, the complex trait of walking onset in the general population and rare genetic disorders that involve delayed or absent walking.

Third, we found genetic correlations between later age at onset of walking and higher cognitive performance and years in education, lower likelihood of ADHD, higher folding index, local gyrification index, cortical surface area and isotropic volume fraction. We note that the direction of the associations was consistent in all four individual cohorts as well as the meta-analysed results, indicating robust findings. In *post hoc* exploratory genetic correlation analyses as well as polygenic score analyses with infant brain imaging measures, we found evidence of significant positive genetic associations between age at onset of walking and infant, childhood, and adult cortical expansion phenotypes.

Research on the timing of milestones in prenatal brain development across humans, primates and other mammals shows that longer duration (more prolonged development) is associated with larger brain volumes, and in particular, enlargement of later developing brain structures^70^. Stemming from our findings reported here, a testable hypothesis is that, to the extent that onset of walking can index a general rate of brain development, the later the onset of walking, the more prolonged is that individual’s time course of brain development, associating later with greater gyrification and higher IQ and educational attainment.

The mechanisms underlying the genetic correlations between later age of walking and higher IQ, more years in education, higher gyrification, folding indices, cortical surface area and water diffusion in the cortex (indicated by the isotropic volume fraction metric) should be explored in future research. The ability to walk requires practice and movement^61^. One behavioural hypothesis is that prior to walking, infants who have longer attention spans and average or lower activity levels may spend more time stationary. Infants with higher activity levels or shorter attention spans may — on average — move about more, thus gaining more practice in movement and muscle strengthening and training, ultimately resulting in earlier walking onset. From a neuroscience perspective, one could speculate that enhanced practice in the first year of life would lead to faster learning, since brain plasticity in the motor system is maximal during this period^71^. Thus, attention and activity levels may influence motor system training in young children. In support of the hypothesis that shorter attention span and higher activity levels would be associated with earlier walking, a recent study of over 25,000 children from MoBa found that the ADHD polygenic score was associated with earlier walking^16^. Further, the ADHD polygenic score was associated with better gross motor skills, such as walking, climbing stairs, and jumping, in 7,498 18-month-old children from the Avon Longitudinal Study of Parents and Children (ALSPAC)^72^. Consistent with these findings, a recent GWAS of infant language found that ADHD is also genetically correlated with better early expressive vocabulary^73^.

A second hypothesis relates to a protracted phase of development across the cortex and, in particular, in the primary motor cortex (M1) over the first year after birth^74^. If the M1 takes longer to develop and build connections in some children, this would put a constraint on when children begin to walk, with later walking associated with higher cognition. In line with this hypothesis, we found that gene sets involved in age at onset of walking are involved in the generation of neurons. Further, we observed that genes associated with age at onset of walking are enriched in the brain between 19 and 24 weeks post conception (Fig. S10). Additionally, the polygenic score predisposing to later onset of walking is associated with larger volumes of brain areas involved in motor control at birth (Fig. 6). Since advantages and costs to early walking might vary based on the individual’s environmental conditions, wide individual differences in the duration of the sensitive period to learn to walk might be the result of the ability of human beings to adapt successfully to their local environment^3,75^.

Current public health policy employs late walking (> 18 months) as a red flag for developmental delay which typically triggers referral for clinical assessment aimed to identify the reason for a departure from the normal range of achievement of this milestone^1^. A better understanding of the entire variation of age at onset of walking can help in more precise intervention planning. In the absence of a deleterious, highly penetrant factor known to cause developmental delay (rare genetic effects and environmental factors leading to an acquired condition such as cerebral palsy, for example), our results suggest that age at onset of walking is part of typical species variation and that late walking may be associated with larger brain volume and surface area, and improved cognitive outcomes. Historical data suggests the majority of late walkers do not have a medically-recognised developmental disorder^9^. Future research should explore whether early walking may also be a useful red flag that may offer early information about likelihood of ADHD or learning difficulties. Further, Mendelian Randomization approaches can be used to understand whether early walking has a causal effect on other domains of neurodevelopment.

Finally, our results from within-family polygenic score analyses suggest that the majority of the signal identified through our GWAS of age at onset of walking captures direct genetic effects and is not confounded with indirect effects such as gene-environment correlation, assortative mating and stochastic effects^47^. Future genetic research should test the relative role of direct and indirect effects for other infant phenotypes once more infant GWAS become available. If true, this offers evidence for an important conceptual point about how genes and environment operate together across the lifespan.

In our study design, we took a comprehensive approach to the phenotype and samples. Relevant samples were searched for using multiple database resources, research council websites and bibliographies. Samples were only included if they had a highly similar phenotype (age at onset of walking in months) and a sample size of greater than 1000 to ensure reliable effect sizes in individual samples. Several forms of evidence, including out-of-sample prediction and measurement harmonisation, supported meta-analysing across our four cohorts. Nevertheless, the potential attrition and participation biases present in population cohorts should be considered in relation to our findings^76,77^. An important limitation of this study is that our meta-analysis only included Western European cohorts, as at the time of conducting the study information on age at onset of walking was not available in other genotyped cohorts that have the power to conduct a GWAS. Extending this investigation to a more diverse population is vital to produce robust findings on the shared biology underlying the onset of bipedal ambulation, especially given the perceived role of different cultural practices in the achievement of this milestone. Some of the phenotypes, such as schizophrenia, major depression and a cross-disorder psychiatric/neurodevelopmental construct did not show significant genetic correlations with age at onset of walking. It remains to be seen, as sample sizes for neuropsychiatric disorder GWAS become larger, whether age at onset of walking will show larger genetic correlations with these phenotypes. It is noted that bipolar disorder shows a significant positive genetic correlation with years in education^53^ and as such, the positive genetic correlation with age at onset of walking is in line with previous findings.

In summary, we identified the first common genetic loci associated with age at onset of walking and report that it is a highly polygenic and heritable phenotype. The results provided candidate causal genes, and implicated genes expressed in the brain, especially between 19 and 24 postconceptional weeks, and genes involved in neurogenesis. The genetic variants identified were highly plausible for a motor phenotype, being previously associated with disorders that disrupt the development of walking and are linked to motor disorders, such as the Brunet-Wagner neurodevelopmental syndrome and the Koolen-de Vries syndrome. Rare mutations of these genes are associated with both motor and cognitive phenotypes (neurodevelopmental disorder and intellectual disability), and for common genetic variation, age at onset of walking was genetically correlated with IQ. We identified a “double hit” haplotype in one of these genes, RBL2, which both decreases gene expression and contains a potentially deleterious missense variant associated with delayed walking. The polygenic score explained 3-5.6% variance in age at onset of walking, the majority of which was direct genetic effects, predicted gross motor skills in an independent cohort and was associated with increased brain volume at birth in regions involved in motor control. Age at onset of walking showed significant genetic correlations with both later outcomes and adult brain structures including IQ, educational attainment, ADHD, BMI, and water diffusion in the cortex, brain gyrification and folding indices.

## Methods

### Samples

The meta-analysis was conducted using data from four birth cohort samples of European ancestry. Full details of the samples are provided in the Supplementary Note A. Analyses were pre-registered on the Open Science Framework (https://doi.org/10.17605/OSF.IO/M2QV3).

#### Lifelines

Lifelines is a multi-generational prospective population-based birth cohort study examining the health and health-related behaviours of 167,729 persons living in the North of the Netherlands. lt employs a broad range of investigative procedures in assessing the biomedical, socio-demographic, behavioural, physical and psychological factors that contribute to the health and disease of the general population, with a special focus on multi-morbidity and complex genetics ^20,78^. Individuals aged 25 to 50 were recruited from the Northern region of the Netherlands between 2006 and 2013 and, during their first study visit, were asked for consent for the study team to approach family members with an invitation to participate. This included any children (≥ 6 months) of cohort members.

Questionnaires about children were answered by parents based on retrospective recollection. The final sample size of Lifelines children with good quality phenotype and genotype data included in the GWAS was 3,415 (1,768 females, 1,647 males).

#### The Norwegian Mother, Father and Child Cohort Study (MoBa)

MoBa is a population-based pregnancy cohort study conducted by the Norwegian Institute of Public Health ^17,18^. Participants were recruited from all over Norway from 1999-2008. The women consented to participation in 41% of the pregnancies. Blood samples were obtained from both parents during pregnancy and from mothers and children (umbilical cord) at birth^79^. The cohort includes approximately 114,500 children, 95,200 mothers and 75,200 fathers. The current study is based on version 12 of the quality-assured data files released for research in January, 2019. The establishment of MoBa and initial data collection was based on a licence from the Norwegian Data Protection Agency and approval from The Regional Committees for Medical and Health Research Ethics. The MoBa cohort is currently regulated by the Norwegian Health Registry Act. The current study was approved by The Regional Committees for Medical and Health Research Ethics (2016/1702). Phenotype information used in this study (year of birth and sex of the participants) was obtained from the Medical Birth Registry (MBRN), a national health registry containing information about all births in Norway.

After post-imputation quality control, the MoBa dataset includes 207,569 individuals of whom 76,577 were children^80^. The final sample size of children from MoBa with European genetic ancestry and good quality genotype and phenotype information included in the GWAS was 58,302 (28,456 females, 29,846 males).

#### MRC National Study for Health and Development (NSHD)

NSHD is a population-based prospective birth cohort study whose participants were infants from single births born in England, Scotland and Wales during one week in March 1946 (N=5,362) to women with husbands^21^. The dataset includes 2,939 genotyped individuals whose DNA was collected at age 53^81^. The sample was roughly representative of the national population of the same age at the time according to a comparison with census data. The final NSHD GWAS sample size including children with available genotype and phenotype was 2,592 (1,295 females, 1,297 males).

#### Netherlands Twin Register (NTR)

The NTR consists of twins, multiples, and their family members. NTR twins and multiples were recruited into the register as new-borns up to a few months after birth starting in 1987^82^. There were no exclusion criteria. Genotyping has been performed on 7,392 individuals for whom there is parent-report data in infancy^83^. For NTR, 6,251 children (3,399 females, 2,852 males) with good quality genotype and available phenotype data were included in the GWAS.

### Phenotype coding

In all samples, individuals whose age at onset of walking was less than 6 months or greater than 36 months were excluded as outside the normative range^1^. MoBa, NSHD and NTR all recorded age at onset of walking in months as an integer variable. In the Lifelines sample, age at first walking was measured as an ordinal scale, using bins of months of age at first walking. These were recorded using the midpoint for each age bin. The upper and lower bins (‘10 months or younger’ and ‘24 months or older’ respectively), were winsorized, re-coding them to 10 and 24 months respectively. The phenotype descriptives for each cohort are reported in the Supplementary Table S1.

### Genotyping, imputation and quality control

Pre- and post-imputation quality control (QC) and imputation procedures were conducted for each cohort following individual study protocols, and according to a Standard Operating Procedure, which was based on the Rapid Imputation for COnsortias PipeLIne (RICOPILI) pipeline^84^. In all the individual cohorts, samples were excluded from the GWAS if they presented excess autosomal heterozygosity, mismatch between self-reported and genetic sex, XXY genotype and other aneuploidies, individual genotyping rate < 90%. Duplicate samples and samples whose genetically determined ancestry did not overlay with the European ancestry cluster based on a reference panel were also excluded. Autosomal SNPs were excluded from the GWAS if they had minor allele frequency (MAF) < 0.5%, Hardy-Weinberg Equilibrium (HWE) exact test at p<1 x 10-6, call-rate < 98%. Full details of the pre- and post-imputation QC are provided in the Supplementary Note A and in Supplementary Table S2.

### Genome-wide association analyses

GCTA^85^ fastGWA^86^ was used for association analyses in MoBa, Lifelines and NTR, and PLINK^87^ 1.9 was used for association analyses in NSHD, where all related individuals (PI-HAT > 0.2) were excluded from the analysis and the sample size was too small to use fastGWA.

Association analyses of the age at onset of walking, as a continuous variable, were carried out using a mixed linear model. Each primary GWAS included the first 10 ancestry principal components as continuous covariates, and sex and genotyping batch as discrete covariates. MoBa included year of birth, and NTR and Lifelines included age at data collection as continuous covariates. NTR included two dummy variables for the genotyping platform as covariates. In MoBa, Lifelines and NTR, where fastGWA was used, a sparse (0.05 cut-off) Genetic Relatedness Matrix was included in the model to account for relatedness in the sample.

GWAS analyses were performed for each of the samples, using the whole dataset and also with the samples stratified by sex.

### GWAS meta-analysis

Summary statistics QC was performed using the R GWASinspector^88^ package on the cohorts’ summary statistics separately. Variants were excluded if they presented invalid or missing value in the chromosome, position, effect and other allele, beta, standard error columns, duplicated alleles, if they were monomorphic (with allele frequency of 0 or 1 and variants with identical alleles), allosomal or mitochondrial, or if they had imputation quality score < 0.8. Results of the summary statistics QC are provided in the Supplementary Information and Supplementary Table S3.

Summary statistics for the four samples were meta-analysed with a standard-error weighted meta-analysis in METAL^24^ on SNPs with MAF > 1%. SNPs were matched between cohorts using rsIDs, which had been assigned according to their chromosome, base-pair positions and alleles based on the 1000 Genomes^29^ reference panel in GWASinspector. Meta-analyses were performed separately for the whole sample, and for sex-stratified samples. Finally, only SNPs for which the minimum sample size was 10,000 were retained for further analyses (6,902,401 variants).

### Fine mapping and functional annotation

In order to identify significant independent SNPs associated with age at onset of walking at each locus at a p-value threshold of p < 5 x 10^-8 89^, we conducted conditional and joint association analyses (COJO)^25^ in GCTA^85^. This analysis conditions on the lead SNP at a locus, and tests for further independent significant SNPs within the same chromosome using a stepwise selection procedure. The MoBa genotype data were used to estimate Linkage Disequilibrium (LD), in line with the COJO guidelines.

Fine mapping, functional annotation and gene-based analyses were carried out in FUMA^30^ (version 1.5.2) and MAGMA^35^ (version 1.08), indicating the list of independent lead SNPs from the COJO analysis. We defined significant SNPs to be independent if they had pairwise LD r^2^ < 0.6. Lead SNPs were defined as having pairwise LD r^2^ < 0.1^90^. Loci were merged if LD blocks distance was < 250 kb.

For gene-mapping in FUMA, SNPs were mapped to genes at a maximum distance of 1 Mb^30^ based on position, eQTL for selected relevant tissues such as the brain, lung, muscles, heart and adipose tissue and chromatin interaction in the brain (see Table S5). Annotation of genes was performed by ANNOVAR within FUMA (date of download 2017-07-17).

A subset of genes prioritised based on mapping using only significant SNP-gene pairs at a False Discovery Rate (FDR) corrected p-value < 0.05, were tested for differential expression in 54 Genotype-Tissue Expression (GTEx) v8^33^ and 11 BrainSpan^32^ tissues and gene-set enrichment using GENE2FUNC in FUMA. The gene set analysis in FUMA was performed to test whether the prioritised genes were over-represented in predefined gene sets obtained from the Molecular Signatures Database^91,92^ (MSigDB) v7.0, WikiPathways^93^ (v 20191010) and GWAS Catalog^94^ (v e0_r2022-11-29) databases, after excluding the MHC region and applying Bonferroni correction for multiple testing.

For MAGMA analyses, the MHC region was excluded and SNPs within 1 kb from a gene were assigned to each gene^90^. The MAGMA gene-based test identified genes associated with age at onset of walking from all 18,766 mapped genes using a Bonferroni correction to define statistical significance (Table S7). The MAGMA gene-property analysis used 53 GTEx v8^33^ and 11 BrainSpan^32^ RNAseq datasets to test tissue specificity of genes associated with age at onset of walking, based on association p-values of all the 18,766 genes mapped in FUMA.

### Colocalization

We used coloc.SuSiE^40^ to identify colocalization of GWAS and eQTL signals, using an LD reference panel of 1,444,196 HapMap3 SNPs with LD calculated in European-ancestry individuals from the UK Biobank^95,96^. Pairs of variants further than 3 cM apart are assumed to have 0 correlation. The eQTL data is from 261 post-mortem bulk RNA-seq samples of human cerebellum^39^. We used a two-sided Wilcoxon rank test to test for differences in *RBL2* expression in the human cortex by genotype for GWAS and eQTL significant SNPs at MAF ∼50% using bulk RNA-seq data of prefrontal cortex from BrainVar^41^ (periods 4-6). Missense variants in the chromosome 16 locus were annotated using the Variant Effect Predictor (VEP)^42^. Protein structure for RBL2 was predicted using AlphaFold^45^. Annotation of p.Tyr210Cys on RBL2 was done using the Genomics 2 Proteins Portal^46^.

### LD score regression

LD score regression (LDSC^22^) was used to calculate h^2^_SNP_ and bivariate genetic correlations, using the 1000 Genomes Phase 3^29^ European ancestry LD scores reference panel. Bivariate genetic correlations were calculated between age at onset of walking and multiple infant, psychiatric, neurodevelopmental and global cortical phenotypes, specifically: birth weight^97^, childhood Body-Mass Index (BMI)^48^, adult BMI^49^, autism^98^, ADHD^50^, educational attainment (EA)^51^, intelligence (IQ)^52^, schizophrenia^99^, general loading for psychiatric disorders^100^, major depression^101^, bipolar disorder^53^, and 13 cortical phenotypes^102^ (see Fig. 5A). Genetic correlation was also calculated between the age at onset of walking in each of the cohorts. Additionally, LDSC was used to calculate h^2^_SNP_ for the female and male meta-GWAS and genetic correlation between the sex-stratified analyses. Statistical significance was evaluated based on 95% confidence intervals as per the pre-registration. Bonferroni-adjusted p-values correcting for 28 multiple testing are reported in the Supplementary Table S12.

As post-hoc analyses, which were not pre-registered, we also used LDSC to test the genetic correlation between age at onset of walking and four other motor phenotypes; self-reported walking pace^103^, clinically ascertained muscle weakness in the pincer grip in elderly people^104^, motor coordination in childhood^105^ and Parkinson’s Disease^106^.

Stratified LDSC^37^ was conducted to obtain estimates of partitioned heritability by functional annotation and cell-type. HapMap3^107^ SNPs (excluding the HLA region) from the meta-GWAS summary statistics weighted by LD score obtained from a European 1000 Genomes^29^ reference panel were used in the regression, as recommended by Finucane and colleagues^37^. To estimate the proportion of genome-wide h^2^_SNP_ attributable to functional categories, we run the stratified LDSC ‘full baseline model’ (described elsewhere^37^) that evaluates whether heritability in a functional category is greater than heritability outside the category. This was tested for 96 functional categories provided by the stratified LDSC developers, including coding, UTRs, promoter and intron annotations from UCSC^108^, genomic annotations for all cell types and fetal cell types only from ENCODE^109^ and the Roadmap Epigenomics Consortium^110^, region conserved in mammals from Lindblad-Toh et al.^111^, FANTOM5 enhancers from Andresson et al.^112^. The p-value for enrichment was adjusted for multiple testing using the Bonferroni method, as in previous similar research^113^. To calculate whether heritability was enriched in specific cell types, we applied stratified LDSC to 53 sets of specifically expressed genes^38^ using multi-tissue gene expression data from the GTEx^33^ project. Bonferroni correction was applied to correct for multiple testing.

### Genomic SEM

A non-pre-registered Genomic SEM^55^ analysis was conducted to test whether the association of the genetic components of age at onset of walking with ADHD remained significant after conditioning for educational attainment. To this aim, we performed a genetic multivariable regression using the same ADHD^50^ and EA^51^ summary statistics that were entered in the LDSC analysis. For ADHD, the sample size was defined as effective sample = 4v*(1-v)*(N cases + N controls) and the sample prevalence as 50%, as indicated by the Genomic SEM developers (https://github.com/GenomicSEM/GenomicSEM/wiki/2.-Important-resources-and-key-information). The summary statistics were munged using HapMap3 SNPs. Both standardised and unstandardised results are reported in Supplementary Fig. S12.

### MiXeR

Univariate causal mixture models were applied using MiXeR^56^, to obtain estimates of polygenicity, defined as the proportion of variants that contribute to 90% of the h^2^_SNP114_. We fitted bivariate models in MiXeR to estimate the genetic overlap that is due to both concordant and discordant SNP effects, between age at onset of walking and eight other phenotypes which had a significant genetic correlation with it (calculated using LDSC). When the summary statistics for the second phenotype in these bivariate analyses came from the case-control GWAS, the N_eff_ was calculated as 4 / (1 / Ncase + 1 / Ncontrol). The MHC (6:26000000-34000000) was excluded from MiXeR analyses due to its complex LD structure. MiXeR v1.3 was used for these analyses, and the data were prepared using scripts developed by the same group (https://github.com/precimed/python_convert).

### Polygenic score analysis

Polygenic scores (PGSs) were calculated using PRS-cs^115,116^; a leave-one-out design was employed whereby additional GWAS meta-analyses were conducted, leaving out one of each of the smaller samples (NSHD, NTR and Lifelines) in turn to be used as a target dataset and meta-analysing the remaining samples as a training dataset for estimation of SNP weights. The MoBa sample comprises most of the overall sample size, and thus could not be used as a target dataset, so a within-MoBa cross-validation was employed. The MoBa dataset was split randomly into five samples of roughly equal size by removing one fifth of the data in turn (with no overlap in these fifths) from the whole dataset to create five new samples, each comprising four fifths of the data. GWASs were then conducted on each of these five new samples, and the summary statistics used for estimation of PGS SNP weights applied to the left-out fifth of the data. This was performed five times, using each of the fifths as target data in turn.

For all leave-one-out PGS analyses, including the within-MoBa design, we derived weights for each chromosome using the 1000 Genomes phase 3 European panel^29^ as a reference for LD, and the following PRS-cs parameters: parameter a and b in the gamma-gamma prior = 1 and 0.5, respectively, global shrinkage parameter phi = 0.01, 1000 MCMC iterations, 500 burn-ins and five as a thinning factor of the Markov chain. PLINK 2.0^117^ was used to compute the PGS in the target sample. The proportion of variance explained by the PGS, scaled so that mean = 0 and SD = 1, was quantified in the NTR cohort by the squared beta-coefficient from a linear regression model between the scaled phenotype and the PGS, including 10 PCs, age, sex and genotyping platform in the model, and was quantified in all other cohorts with adjusted R^2^ of the linear regression between the scaled phenotype regressed on 10 PCs and the genotype batch and the PGS.

### Within- and between-family polygenic score analysis

Within- and between-family analyses were performed using the NTR cohort dataset. The method is described in^47^, and scripts were used from ^118^ (https://github.com/PerlineDemange/GeneticNurtureNonCog/).

A PGS was generated from a meta-analysis of the MoBa, Lifelines and NSHD GWAS (calculated as above), and the predictive power of this PGS was quantified in the whole NTR sample using the above method. We used a random intercept mixed-effects linear model in R using the dizygotic twins-only subsample of NTR (N = 2,586 individuals in 1,293 twin pairs), after ensuring that a mixed-effects model was justified by calculating a bootstrapped intraclass correlation (ICC). PGS entered into the model were first scaled to mean = 0 and SD = 1. Within-family PGS effects were calculated by subtracting the family mean PGS from each individual PGS. Between-family effects were modelled using the mean family PGS. The linear model included age, sex, the first 10 PCs and a genotyping platform dummy variable as covariates. The within- and between-family standardised regression coefficients were compared by a χ^2^ test.

### Polygenic score in the Developing Human Connectome Project

#### Genetic data

Infant saliva DNA was genotyped for SNPs genome-wide on the Illumina Infinium Omni5-4 array and standard quality control was performed. The dataset was imputed to the Haplotype Reference Consortium reference panel^119^ on the Michigan Imputation Server. The imputed data were used to compute an age at onset of walking PGS for each of the 264 unrelated European infants using summary statistics from the age at onset of walking meta-GWAS and the PRS-cs software^115^, as previously described.

#### Imaging data: Acquisition and processing

T2-weighted MRI data were acquired at term equivalent age (median postmenstrual age = 41.9 weeks) as part of the Developing Human Connectome Project (dHCP^58^). T2 images were registered to the 40-week dHCP neonatal atlas (https://brain-development.org/brain-atlases/atlases-from-the-dhcp-project/)^120^ via an age-matched intermediate using Symmetric Diffeomorphic Image Registration, implemented using Advanced Neuroimaging Tools (ANTs)^121,122^ as a measure of individual variation in brain volume, the log-Jacobian determinant images were calculated by applying ANTs algorithms to the non-linear transformation deformation tensor fields. Log-Jacobian maps were then smoothed using a 3 mm full-width half-maximum Gaussian filter and down sampled to 1 mm isotropic resolution (to increase computational efficiency). A 4D volume was created by merging the 1 mm log-Jacobian maps across all subjects (N = 264), then subsequently used as the input to the randomise algorithm (described below).

#### Imaging data: Tensor based morphometry

The randomise function, part of the FMRIB Software Library (FSL)^23,123^ was used to apply a general linear model, including gestational age, postmenstrual age at scan, sex, weight-z-score and 10 ancestral PCs as covariates. Threshold-Free Cluster Enhancement (TFCE) and Family-Wise Error (FWE) rate were applied to correct for multiple comparisons between voxels. Significant areas were identified with permutation testing using 5,000 random permutations. We show results at a significance level of p < 0.05 in the FWE-corrected contrast.

## Data Availability

All data produced in the present study are available upon reasonable request to the authors

## Acknowledgements

This work was funded by the Simons Foundation to AR (Award ID: 724306). AHollowell was supported by the Economic and Social Research Council (ES/P000592/1). AHavdahl, LH and EC were supported by the South-Eastern Norway Regional Health Authority (#2020022, #2022083, and #2021045, respectively). AHavdahl and EC were supported by the Research Council of Norway (RCN) (#336085 and #274611, respectively). AW was supported by the UK Medical Research Council (MC_UU_00019/1). SW and TA received support from the Medical Research Council Centre for Neurodevelopmental Disorders, King’s College London (MR/N026063/1). The Developing Human Connectome Project was funded by the European Research Council under the European Union Seventh Framework Programme (FP/2007– 2013)/ERC Grant Agreement No. 319456. HC is funded by a National Institute for Health Research (NIHR), Academic Clinical Lectureship at King’s College London. OAA received support by KG Jebsen Stiftelsen, Research Council of Norway (#324499, #324252, #223273); EU H2020 RIA grant# #964874 REALMENT. MB was supported by the ERC WELL-BEING 771057. DIB received support by the KNAW Academy Professor Award (PAH/6635). TA was supported by an MRC Clinician Scientist Fellowship (MR/P008712/1, MR/Y009665/1) and a Transition Support Award (MR/V036874/1). DE received support by the European Research Counci (FP/20072013). MJ received support from MRC PG (MR/T003057/1). This work was supported by grants from the NIMH (R01 MH129751 and U01 MH122681 to SJS) and HDR-UK to SJS.

The Norwegian Mother, Father and Child Cohort Study is supported by the Norwegian Ministry of Health and Care Services, and the Ministry of Education and Research. We are grateful to all the participating families in Norway who take part in this on-going cohort study. We thank the Norwegian Institute of Public Health (NIPHI) for generating high-quality genomic data. This research is part of the HARVEST collaboration supported by the RCN (grant #229624). For providing genotype data, we also thank the NORMENT Centre (funded by the RCN (#223273), South-Eastern Norway Regional Health Authority (SENRHA) and Stiftelsen Kristian Gerhard Jebsen), in collaboration with deCODE Genetics, and the Center for Diabetes Research at the University of Bergen (funded by the ERC AdG project SELECTionPREDISPOSED, Stiftelsen Kristian Gerhard Jebsen, Trond Mohn Foundation, the RCN, the Novo Nordisk Foundation, the University of Bergen, and the Western Norway Regional Health Authority). This work was performed on the TSD (Tjeneste for Sensitive Data) facilities, owned by the University of Oslo, operated and developed by the TSD service group at the University of Oslo, IT-Department (USIT). The computations were performed on resources provided by Sigma2 - the National Infrastructure for High Performance Computing and Data Storage in Norway.

The Lifelines initiative has been made possible by subsidy from the Dutch Ministry of Health, Welfare and Sport, the Dutch Ministry of Economic Affairs, the University Medical Center Groningen (UMCG), Groningen University and the Provinces in the North of the Netherlands (Drenthe, Friesland, Groningen). The authors wish to acknowledge the services of the Lifelines Cohort Study, the contributing research centres delivering data to Lifelines, and all the study participants.

The MRC National Survey of Health and Development is funded by the UK Medical Research Council [MC_UU_00019/1]. We also thank the study participants for their continuing participation in the National Study of Health and Development and also the study members from the MRC NSHD for their lifelong commitment to the study. Data used in this publication are available to bona fide researchers upon request to the NSHD Data Sharing Committee via a standard application procedure. Further details can be found at http://www.nshd.mrc.ac.uk/data. doi: 10.5522/NSHD/Q101.

The Netherlands Twin Register acknowledges funding from the Netherlands Organization for Scientific research (NWO), including NWO-Grants NWO/SPI 56-464-14192 and 480-15-001/674: Netherlands Twin Registry Repository and the Biobanking and Biomolecular Resources Research Infrastructure (BBMRI–NL, 184.021.007 and 184.033.111); Amsterdam Public Health (APH) and Neuroscience Campus Amsterdam (NCA); the European Community 7th Framework Program (FP7/2007-2013): ENGAGE (HEALTH-F4-2007-201413) and ACTION (9602768) and European Research Council (ERC-230374). We also acknowledge The Rutgers University Cell and DNA Repository cooperative agreement (NIMH U24 MH068457-06); the Collaborative Study of the Genetics of DZ twinning (NIH R01D0042157-01A1); the Developmental Study of Attention Problems in Young Twins (NIMH, RO1 MH58799-03); Major depression: stage 1 genome-wide association in population-based samples (MH081802); Determinants of Adolescent Exercise Behavior (NIDDK R01 DK092127-04); Grand Opportunity grants Integration of Genomics and Transcriptomics (NIMH 1RC2MH089951-01) and Developmental Trajectories of Psychopathology (NIMH 1RC2 MH089995); and the Avera Institute for Human Genetics, Sioux Falls, South Dakota (USA). We also would like to thank the participants and parents for their voluntary participation in research project of The Netherlands Twin Register.

We also acknowledge the Developing Human Connectome Project.

We would like to thank all the families who contributed with their data. We also would like to thank Sarah Medland, Longda Jiang, Espen Hagen, Guy Hindley, Jan Yang and Ruilei Ma for useful advice at various stages of the research.

## Conflicts of interest

OAA is Consultant to cortechs.ai, speakers honoraria from Janssen, Lundbeck, Sunovion. SJS receives research funding from BioMarin Pharmaceutical.

## Notes

### Competing Interest Statement

Ole A. Andreassen is a consultant to cortechs.ai, speakers honoraria from Janssen, Lundbeck, Sunovion. Stephan J. Sanders receives research funding from BioMarin Pharmaceutical.

### Funding Statement

Simons Foundation.

Economic and Social Research Council.

South-Eastern Norway Regional Health Authority.

Research Council of Norway (RCN).

UK Medical Research Council.

Medical Research Council Centre for Neurodevelopmental Disorders, Kings College London.

European Research Council.

National Institute for Health Research (NIHR).

KG Jebsen Stiftelsen.

National Institute of Mental Health.

Health Data Research UK.

Research Council of Norway.

Stiftelsen Kristian Gerhard Jebsen.

Trond Mohn Foundation.

The Novo Nordisk Foundation.

The University of Bergen.

The Western Norway Regional Health Authority.

The Dutch Ministry of Health, Welfare and Sport.

The Dutch Ministry of Economic Affairs.

The University Medical Center Groningen (UMCG).

The Groningen University.

The Provinces in the North of the Netherlands (Drenthe, Friesland, Groningen).

Netherlands Organization for Scientific research.

The Biobanking and Biomolecular Resources Research Infrastructure.

Amsterdam Public Health (APH).

Neuroscience Campus Amsterdam (NCA).

European Community 7th Framework Program.

The Rutgers University Cell and DNA Repository cooperative agreement.

The National Institute of Health (NIH).

The Rutgers University Cell and DNA Repository cooperative agreement.

The Collaborative Study of the Genetics of DZ twinning.

The Developmental Study of Attention Problems in Young Twins.

Major depression: stage 1 genome-wide association in population-based samples.

Determinants of Adolescent Exercise Behavior.

Grand Opportunity grants Integration of Genomics and Transcriptomics.

Developmental Trajectories of Psychopathology.

The Avera Institute for Human Genetics, Sioux Falls, South Dakota (USA).

### Author Declarations

The Departmental Ethics Committee of the Psychological Science Department of Birkbeck, University of London gave ethical approval for this work.

