## Supplementary Notes and Figures for "Genome-wide association meta-analysis of age at onset of walking"

### A. Cohort Descriptions

1) MoBa

The Norwegian Mother, Father and Child Cohort Study (MoBa)[1,2] is a population-based pregnancy cohort from across Norway consisting of approximately 114,500 children, 95,200 mothers and 75,200 fathers[3]. Participants were recruited through hospitals, firstly in Bergen starting in 1999, and then expanding to 50 of 52 Norway's hospitals with maternity units. All pregnant women were eligible for inclusion and 41% accepted of the 277,702 invited between 1999 and 2008 (N = 112,908). About 1,900 twin pairs were born in the MoBa study, and about 16,400 women participated in MoBa with multiple different pregnancies[2]. Due to the wide inclusion criteria, there are numerous related families within the MoBa sample giving it a complex relatedness structure[3].

*Phenotype*

The current study is based on version 12 of the quality-assured questionnaire data files released for research in January 2019. The phenotype and genetic data analysed were prepared on the 5th of December 2022. Information on gross motor skills was collected by parental questionnaire at 18 months and 36 months after birth. The parental questionnaire administered when children were 18 months asked 'How many months was your child when he/she took his/her first steps unaided (questionnaire version A and B; variable EE400) (N = 17,488) or "Can your child walk unaided? If yes, how old was your child when he/she could first walk unaided?" (questionnaire version C, D and E; variable EE987) (N = 56,715). If data was unavailable for a participant, including if independent walking had not yet been attained at 18 months, responses from a questionnaire administered when children were 36 months was used, being "How many months old was your child when he/she took his/her first steps unaided?" (all questionnaire versions – no edits made; variable GG27) (N = 5,594).

Phenotype descriptive statistics (genotyped participants only) for MoBa are provided in the Supplementary Table S1.

*Genotype and Quality Control (QC)*

Blood samples from infants were taken from the umbilical cord at birth[4]. Genotyping of the samples took place at different times as part of different projects over several years. Including mothers, fathers and children, 238,001 samples were genotyped in 24 batches (two of which were split into sub-batches). The arrays used for the different batches include: Illumina HumanCoreExome12v1.1, Illumina HumanCoreExome24v1.0, Illumina HumanOmniExpress24v1.0, Illumina HumanOmniExpress24v1.2, Illumina Global Screening Array MD v1.0, Illumina Global Screening Array MD v3.0., and Illumina InfiniumOmniExpress-24v1.2.

Of the 238,001 samples genotyped in 26 batches, 235,412 were successfully genotyped and 234,505 of those were retained in the cohort after excluding those who had not withdrawn consent. Based on the principal components analysis, approximately 95% of the sample were identified as having European ancestry. The publicly available European Haplotype Reference Consortium (HRC)[5] release 1.1 was used as the reference panel for phasing and imputation. Full details of the genotyping and QC pipeline, including the list of researchers involved in this process, can be found here: [3]. After post-imputation quality control, the dataset included 207,569 individuals (including 76,577 children) and 6,981,748 SNPs. Further post-imputation QC in MoBa as well as the primary and sex stratified Genome-Wide Association Study (GWAS) were conducted by Anja Hollowell and Anna Gui for children who had phenotype data concerning motor or behavioural traits (N = 67,407). At this stage, individuals were further excluded if they had genotyping rate < 95%. SNPs were excluded if they had minor allele frequency (MAF) < 0.5%, failed Hardy Weinberg Equilibrium (HWE) exact test at $p < 1 \times 10^{-6}$, or had call-rate < 98%. In addition, to exclude potential duplicates, for each of the 33 pairs samples that were genetically identical (P(identity by descent or IBD=0)<0.1 & P(IBD=1)<0.1, PI-HAT>0.8), and also identical across 12 infant phenotype measures (age at onset of walking, two other motor measures, and 9 behavioural measures), one of the pair was excluded from the data at random.

The current GWAS included children with complete information on age at onset of walking, covariates (sex, year of birth, genotyping batch, imputation batch and the first 10 principal components, PCs), and genotype data (N = 58,302). Sex-stratified GWAS analyses were performed on 29,846 males, and 28,456 females, including year of birth, genotyping batch, imputation batch and the first 10 PCs as covariates in the model. The QQ-plot for the MoBa GWAS is shown in the Supplementary Fig. S1.

*Additional checks for inflation in the MoBa samples*

Since the Genomic Control (GC) factor lambda ($\lambda_{GC}$) for the MoBa GWAS was $\lambda_{GC} = 1.228$, we ran some additional checks to verify whether the inflation observed in the QQ-plot (Supplementary Fig. S1) was due to bias linked to the sample ancestry and relatedness, phenotypic outliers, covariate-phenotype interactions or variant allele frequencies, variant LD patterns or more likely reflected polygenicity. These checks are summarised in Table SM1 below.

The MoBa sample only included individuals of European ancestry and the GWAS model included 10 PCs to correct for residual population stratification. Additionally, the $\lambda_{GC}$ for a GWAS on a different infant motor phenotype collected at the same age from a largely overlapping sample (N = 55,227), namely fine motor skills from parental report at 18 months was 1.040. This result strongly suggested that the observed inflation was not due to population stratification but was linked to the age at onset of walking phenotype specifically. Additionally, the QQ-plot by allele frequency (see Fig. S1) showed that the observed pattern was not changed by further filtering by allele frequency. Thus, we performed additional GWAS analyses and computed the $\lambda_{GC}$, to see whether inflation was caused by particular characteristics of the data.

To check whether the observed inflation pattern in the QQ-plot was due to the presence of sample outliers, we randomly split the sample in halves and computed the $\lambda_{GC}$ for both samples (N = 29,151 each, $\lambda_{GC}$ = 1.117 and 1.123). We then repeated this operation and computed the lambda GC for the new two-halves (N = 29,151 each, $\lambda_{GC}$ = 1.112 and 1.131 respectively). The similarity in lambdas across the two halves suggest there is not a specific individual/subset of individuals driving the high total sample lambda. The lower lambdas for the half-samples is consistent with the idea that inflation reflected polygenicity, as smaller sample sizes reduce power therefore the Chi-squared statistics are lower.

The age at onset of walking phenotype for MoBa ranged from 6 months to 33 months and was not normally distributed (skew=0.91). To verify whether inflation could be caused by the presence of phenotype outliers, we excluded individuals who walked later than 25 months of age, which corresponded to the latest age at onset of walking reported in the other cohorts (as summarised in the Supplementary Table S1). The $\lambda_{GC}$ for this GWAS was 1.230, suggesting that extreme phenotype outliers were not causing genomic inflation in the primary MoBa GWAS. Further, to exclude that this effect was caused by skewness in the phenotype distribution, we excluded all phenotype values above 3 SD from the mean of the whole sample, that is those who learnt to walk later than 18 months. Again, we found the $\lambda_{GC}$ = 1.233, indicating that inflation was not due to the presence of phenotype outliers.

We then tested whether the observed pattern of inflation in the MoBa GWAS was due to the inclusion of specific covariates in the GWAS model. When excluding genotyping and imputation batch covariates, the $\lambda_{GC}$ was 1.228; when excluding the year of birth covariate, the $\lambda_{GC}$ was 1.226; when excluding year of birth and batches from the model, leaving only sex and the 10 PCs as covariates, the $\lambda_{GC}$ was 1.225. These analyses suggested inflation was not caused by the covariates included in the primary MoBa GWAS.

Since one of the most significant signals in the MoBa GWAS was on Chromosome 6 in the HLA region, we conducted an additional check by removing chromosome 6 from the GWAS to verify whether the LD pattern in that part of the genome caused inflation. However, $\lambda_{GC}$ for the GWAS without chromosome 6 was 1.222, similar to the $\lambda_{GC}$ for the primary MoBa GWAS.

Last, the LD Score Regression (LDSC) intercept for the primary MoBa GWAS was 1.008 (0.008), confirming that the inflation observed in the MoBa GWAS QQ-plot was likely due to polygenicity of the phenotype rather than confounding factors such as relatedness and population stratification[6]. Indeed, our MiXeR[7] analyses confirmed that the age at onset of walking phenotype is highly polygenic, with 11.9K variants explaining 90% of the SNP heritability. Other highly polygenic traits had values of $\lambda_{GC}$ higher than 1.2. For example, for ADHD[8] the MiXeR polygenicity estimate was 7.3K variants explaining 90% of the SNP heritability and $\lambda_{GC}$ was 1.307, for bipolar disorder[9] the MiXeR polygenicity estimate was 8.6K and $\lambda_{GC}$ was 1.38). Moreover, inflation was observed for GWAS of many polygenic quantitative traits on the UK BioBank dataset, which is an ancestrally homogeneous sample where all related individuals had been excluded (for example, sitting height showed a $\lambda_{GC}$ of 1.62 for SNPs with MAF > 0.5 and $\lambda_{GC}$ = 4.034 when including all SNPs with MAF > 0.01, https://pheweb.org/UKB-Neale/pheno/20015 , or heel bone mineral density with a $\lambda_{GC}$ of 1.20 for MAF > 0.5 and $\lambda_{GC}$ = 1.605 for MAF > 0.01 https://pheweb.org/UKB-Neale/pheno/4125#qq , see the NealeLab blog for a discussion regarding inflation for polygenic traits: https://www.nealelab.is/blog/2017/9/11/details-and-considerations-of-the-uk-biobank-gwas ). Further, a similar QQ plot showing a "step-like" line linked to LD was observed in the recent GWAS of educational attainment[10] for SNPs with MAF > 5% (Extended Fig.1 a, $\lambda_{GC}$ = 3.55).

Based on these additional checks, and because all other samples had $\lambda_{GC}$ below the 1.10 recommended threshold[11], we decided not to apply automatic correction for genomic control based on the $\lambda_{GC}$ in the GWAS meta-analysis, to avoid removing meaningful signal.

Table SM1. Summary of the inflation factors for all the analyses performed to examine possible reasons for the high genomic inflation.

| Possible reason | Performed analysis | Lambda GC |
|---|---|---|
| The inflation is due to population stratification or relatedness in the sample | GWAS of fine motor skills at 18 months in a largely overlapping sample | 1.040 |
| The inflation is due to a specific individual/subset of individual outliers | Randomly split the sample into two halves.<br><br>Repeat random split into two halves. | 1.117<br>1.123<br>1.112<br>1.131 |
| The inflation is due to phenotype outliers | Exclude individuals who walked later than 25 months (upper limit in NSHD and NTR cohorts)<br><br>Exclude individuals with phenotype values exceeding 3 standard deviations from the sample mean | 1.230<br><br>1.233 |

| | | |
|---|---|---|
| The inflation is due to the interaction between covariates and the phenotype | Exclude genotyping and imputation batch covariates | 1.228 |
| | Exclude the year of birth covariate | 1.226 |
| | Exclude the year of birth, genotyping and imputation batch covariates | 1.225 |
| The inflation is due to the LD pattern in the HLA region | Exclude chromosome 6 from the GWAS | 1.222 |

*Ethics*

MoBa and the related data collection was authorised by a licence from the Norwegian Data Protection Agency and an approval from the The Regional Committees for Medical and Health Research Ethics (REK). MoBa is regulated by the Norwegian Health Registry Act. The current study was approved by REK (2016/1702).

2) Netherlands Twin Register (NTR)

The Netherlands Twin Register (NTR)[20,21] is a population-based cohort of over 200,000 people from across the Netherlands. It consists of twin-families, i.e. twins, their parents, spouses and siblings aged between 0 and 99 years at recruitment. NTR started around 1987 with new–born twins and adolescent and adult twins. Full details have been reported previously[22]. The current sample includes monozygotic and dizygotic twins and triplets who were recruited shortly after birth.

*Phenotype*

Information on age of walking without support was obtained from NTR survey YS-2, which was mailed to the parents after the second birthday of the children (mean age 2.34 years, SD 0.25). The parents (usually the mother of twins) were asked “With how many months could your twin walk without support for the first time? in half months”. With the first survey that

was sent to parents, typically a few weeks/months after registration we informed them that they would be asked for this information in the next survey and included an example of the answer sheet. The procedure for collection of the information was validated in a large study[18,19]. The answers on age of walking without support ranged from 9.5 to 21 months (mean 15.13, SD 2.36, median 15, interquartile range 3).
Phenotype descriptive statistics for the NTR sample included in the present GWAS are provided in the Supplementary Table S1.

*Genotyping and QC*

Buccal cells or blood samples for DNA isolation were collected in multiple NTR projects, thus genotype data was obtained from participants at different time points. Genotyping in NTR was done on multiple platforms over time, including Perlegen-Affymetrix, Affymetrix 6.0, Affymetrix Axiom, Illumina Human Quad Bead 660, Illumina Omni 1M and Illumina GSA, in total on 29,943 individuals. On each platform, genotyping was performed following manufacturers protocols, using the appropriate calling software. For each genotype platform, samples were removed if DNA sex did not match the expected phenotype, if the PLINK heterozygosity F statistic was $< -0.10$ or $> 0.10$, or if the genotyping call rate was $<0.90$. DNA IBD estimations were made for each platform and samples were removed if they did not match the expected relationships (PLINK[19], KING[23]). SNPs were removed if the MAF $< 1\times10^{-6}$, if the HWE p-value $< 1\times10^{-6}$, and if call rate $< 0.95$.

Subsequently, for each platform, genotype data were aligned with 1000 Genomes reference[15] panel using the HRC and 1000 Genomes checking tool, which tests and filters for SNPs with allele frequency differences larger than 0.20 as compared to the CEU population, palindromic SNPs and DNA strand and position issues. The data of the 6 platforms were then merged into a single data set keeping all QC-ed SNPs of each platform. For each individual, only one platform was chosen. Based on the ~10.8K SNPs that all platforms have in common, IBD state was then estimated for all individual pairs using the PLINK and KING programs. These estimates were then compared to the expected familial relations, and any individuals whose genetic relatedness estimate did not match their expected familial relations were removed.

Principal components were calculated for each platform after genotype QC, with additional LD SNP pruning (VIF <=2), in order to exclude CEU population outliers, based on 1000 genomes population projection with the SMARTPCA software[24]. Then, per platform, data were phased using EAGLE[25] and then imputed to 1000 Genomes[15] with Minimac-3[26]. Post-imputation, the resulting separate platform VCFs were merged into a single VCF file per chromosome using BCFTOOLs for all SNPs present on each of the 6 platforms.

The current GWAS included children with complete information on age at onset of walking without support, covariates (age at parents' report, sex and platform), and genotype data (N = 6,251, born between 1986-2016). Sex-stratified analyses were performed in 2,852 males, and 3,399 females. In this case 2 platform covariates, minus 1 to avoid collinearity were added, depending on the individuals with phenotypes used in the study. The described QC steps and GWAS were conducted by Jouke-Jan Hottenga and Veronika Odintsova. The QQ-plot from the primary NTR GWAS is shown on Supplementary Fig. S2.

*Ethics*

Informed consent was obtained from parents or guardians. The study was approved by the Central Ethics Committee on Research Involving Human Subjects of the VU University Medical Centre, Amsterdam, an Institutional Review Board certified by the U.S. Office of Human Research Protections (IRB number IRB00002991 under Federal-wide Assurance-FWA00017598; IRB/institute codes, NTR 03-180).

3) The Lifelines Cohort Study (Lifelines)

Lifelines is a multi-disciplinary prospective population-based cohort study examining in a unique three-generation design the health and health-related behaviours of 167,729 persons living in the North of the Netherlands. It employs a broad range of investigative procedures in assessing the biomedical, socio-demographic, behavioural, physical and psychological factors which contribute to the health and disease of the general population, with a special focus on multi-morbidity and complex genetics[12,13]. The study was established in 2006 and asked all the general medical practitioners operating in the provinces of Friesland, Groningen and

Drenthe to invite their patients aged between 25 and 50 to take part in the study unless the patient met one of five exclusion criteria (as determined by the practitioner): a) having a severe psychiatric or physical illness (such that the individual was not fully capable to make rational decisions), b) life expectancy of less than 5 years, c) being unable to complete a Dutch language questionnaire, d) a lack of ability in the Dutch language, or e) not being able to visit their medical practitioner (http://wiki-lifelines.web.rug.nl/doku.php?id=cohort). Those who were interested in participating in the study were required to give written consent and then complete a baseline questionnaire. Parents of children could also volunteer their children for participation. Residents of the relevant provinces were also able to join the study through the study website[13,14].

*Phenotype*

On admission, Lifelines study participants completed questionnaires which, for parents of children taking part in the study, included questions on the child's development and behaviour. During the first assessment of participants, known as the baseline assessment, parents were provided with questionnaire 1A which, amongst other things, asked "When did your child start walking (3-5 steps without help)?" (N ~12,000). We refer to Supplementary Table S1 for information about the phenotype coding and the descriptives for age at onset of walking phenotype for the Lifelines sample included in the present GWAS as providing both phenotype and good quality genotype data (see below).

*Genotype and QC*

During a visit to one of the twelve research sites set up in their local areas, participants provided blood samples. The blood samples were analysed at the Lifelines laboratory and genetic information extracted for sub-samples of Lifelines.

Genetic data was collected as part of two subsequent studies of which only the second collected data from children. This study was called the University Medical Centre in Groningen (UMCG) Genetics Lifelines Initiative (UGLI) as part of which genotyped 38,030 Lifelines participants.

The UGLI data from approximately 38,000 participants (which includes all the genotyped children) was genotyped using the Infinium Global Screening Array (GSA) MultiEthnic Disease Version 1.0. This additional assessment selected 38,030 Lifelines participants with the criteria that they were of European ancestry and an adequate DNA sample for the participant was available. The UGLI participants' data was genotyped in 31 different batches over a period of 5 months in two laboratories. The quality control steps carried out on the entire UGLI sample are described in the UGLI 1.1 Quality Control report and summarised in the Supplementary Table S2.

Following these QC procedures on the total (adult and child) UGLI dataset, 36,339 samples and 571,420 markers remained on autosomal and X chromosomes. Imputation was carried out using the Sanger imputation service and the HRC panel[5]. These pre-processing steps were curated by Serena Sanna, Patrick Deleen, Raul Aguirre-Gamboa, Gerben van der Vries, Ilja Nolte and Esteban Lopera-Maya.

Further post-imputation QC in Lifelines and the GWAS were conducted by Anja Hollowell and Anna Gui. SNPs were filtered based on MAF (< 0.5%), HWE exact test ($p < 1 \times 10^{-6}$) and had call-rate (< 98%). Individuals were further excluded if they had genotyping rate < 95% and if they did not provide phenotype information about early motor and temperament traits, leaving a sample of 3,483 individuals.

X-chromosome data were available only for the Lifelines dataset and was used for sex checks. Post-imputation QC removal of variants and individuals was conducted on X-chromosome data using the same criteria as described above. The pseudo-autosomal region of the X chromosome was removed during QC. X chromosome data were used to identify any individuals for whom there was a discrepancy between phenotypic and genetic sex (N < 10), who were then excluded from analyses.

To ensure each sample was ancestrally homogeneous and aligned with a European reference panel, PCs were calculated, and pairwise plots of PCs along with 1000 Genomes[15] phase 3 reference panel data were visually inspected to identify any individuals to be excluded from analyses. Less than ten individuals for whom either PC1 or PC2 fell outside of 4 standard deviations (SDs) of the centroid of the 1000 Genomes mean were removed.

Finally, IBD was calculated using a pruned set of SNPs and all samples with PI-HAT >0.2 were examined using a relatedness plot[16]. For each pair of samples deemed to be genetically identical (P(IBD=0) < 0.1 & P(IBD=1) < 0.1, PI-HAT > 0.8), phenotype data were inspected. For those genetically identical pairs that also had identical phenotype across three measures (age at first steps and two early temperament measures), one sample of the pair was excluded at random as a plausible duplicate. As a result of this process, 10 samples were removed.

From the UGLI dataset, 3,415 individuals (N males = 1,647, N females = 1,768) with phenotype information on age at onset of walking and covariates (sex, child's age at parental report, batch and the first 10 PCs) were included in the primary and sex stratified GWAS analyses for the present study. The Supplementary Fig. S3 shows the QQ-plot from the primary Lifelines GWAS of age at onset of walking.

*Ethics*

Participants in Lifelines gave written consent prior to physical examination. The study is conducted according to the principles of the Declaration of Helsinki and in accordance with the UMCG research code and is approved by the medical ethical committee of UMCG (document number METC UMCG METc 2007/152).

4) MRC National Study for Health and Development (NSHD)

The NSHD[17,18] is a population maternity cohort where all women who gave birth in a single week in March 1946 in England, Wales or Scotland were targeted for enrolment. From these 16,695 births, 13,687 were participants in the first stage of the study. Later stages of the study only included subsets of this initial sample.

*Phenotype*

The assessment when the children in the cohort were aged 1 to 4 years between 1947 and 1950 included 5,362 participants who were selected with the criteria that the births were single births, and the mothers were married. Data was collected by a health visitor in the mother's home at age 2 years (N = 4,689). One of the items from the questionnaire administered at this time-point was "how many months old was baby when he walked several steps without support".

A follow up assessment when the birth cohort were aged 53 years, where 2,592 participants (N males = 1,297, N females = 1,295) with phenotype information on the date at age at the onset of walking were included in the GWAS analyses. Supplementary Table S1 provides the descriptives for age at onset of walking phenotype for the entire NSHD sample and by sex.

*Genotype and QC*

Blood samples were collected. Illumina DrugDev Consortium Array was used to genotype the biological samples and QC analyses were performed using PLINK[19] 1.9. Sample QC removed individuals with call rates <95%, extreme heterozygosity ($\mu \pm 3$ SDs), sex mismatches, relatedness and duplicates (PI-HAT>0.2), and PC outliers. All participants were of European ancestry. Genotyped SNPs were excluded on the basis of the following parameters: call rate < 95%, MAF < 0.01 or HWE p-value < $1 \times 10^{-4}$. The genotyped SNPs were used to impute information on missing common variants using the HRC[5] v1.1 reference panel, accessed via the Michigan Imputation Server. QC of imputed data led to SNPs being excluded with INFO score < 0.3, MAF < 0.5%, call rate > 98%, HWE p-value < $1 \times 10^{-6}$. Pre- and post-imputation QC was carried out by Valerie Kuan.

The primary and sex stratified GWAS analyses were conducted by Andrew Wong. Sex and the first 10 ancestry PCs were added as covariates in the primary GWAS model (see Supplementary Fig. S4 for the QQ-plot). Only the first 10 PCs were used as covariates in the sex stratified GWAS model.

*Ethics*

The collection of blood samples and DNA information from the participants was approved by ethical approval reference MREC no. 98/2/121.

**B. Study Ethics**

This study and the related secondary data analysis were approved by the Departmental Ethics Committee of the Psychological Science Department of Birkbeck, University of London on 27th October 2020 (reference number 2021007).

**C. Genetic correlation between cohorts**

The SNP-based heritability ($h^2_{SNP}$) estimated using LDSC[6] for the MoBa sample was $h^2_{SNP}$ = 25.11% (SE = 1.34%) and for the NTR sample $h^2_{SNP}$ = 19.09% (SE = 7.42%). Lower $h^2_{SNP}$ estimates and larger standard errors were obtained for the smaller samples, namely Lifelines ($h^2_{SNP}$ = 9.52%, SE = 12.98%) and NSHD ($h^2_{SNP}$ = -3.02%, SE = 17.17%). Genetic correlation ($r_g$) between MoBa and NTR was $r_g$ = 0.89 (SE = 0.17, p = 1.803 x 10-7) and between NTR and Lifelines was $r_g$ = 0.46 (SE = 0.55, p = 0.404). Other genetic correlations were out of bound (MoBa-Lifelines $r_g$ = 1.168, SE = 0.72, p = 0.103) or non-estimable (MoBa-NHSD $r_g$ , Lifelines-NHSD $r_g$ and NTR-NHSD $r_g$ = nan) due to low power, as indicated by the large SNP-based heritability standard errors obtained for the smaller cohorts.

### Supplementary Figures

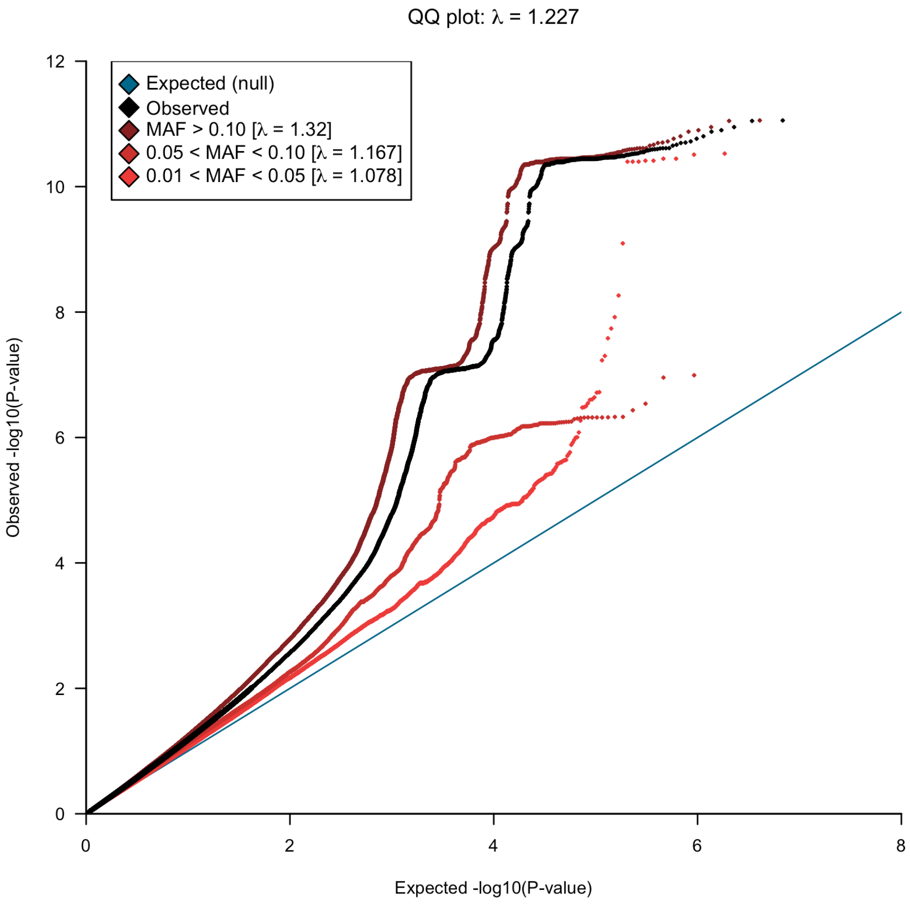


Supplementary Fig. S1. Quantile-quantile plot of the observed $-\log_{10}$[p-values] from MoBa GWAS for age at onset of walking against the expected $-\log_{10}$[two-sided p-value] (blue line). Results are presented by allele frequency, brown dots representing signals for the SNPs with minor allele frequency (MAF) > 10%, red for SNPs with MAF between 5 and 10%, orange circles for SNPs with MAF between 1 and 5% and black circles for all SNPs included in the GWAS (MAF > 1%).

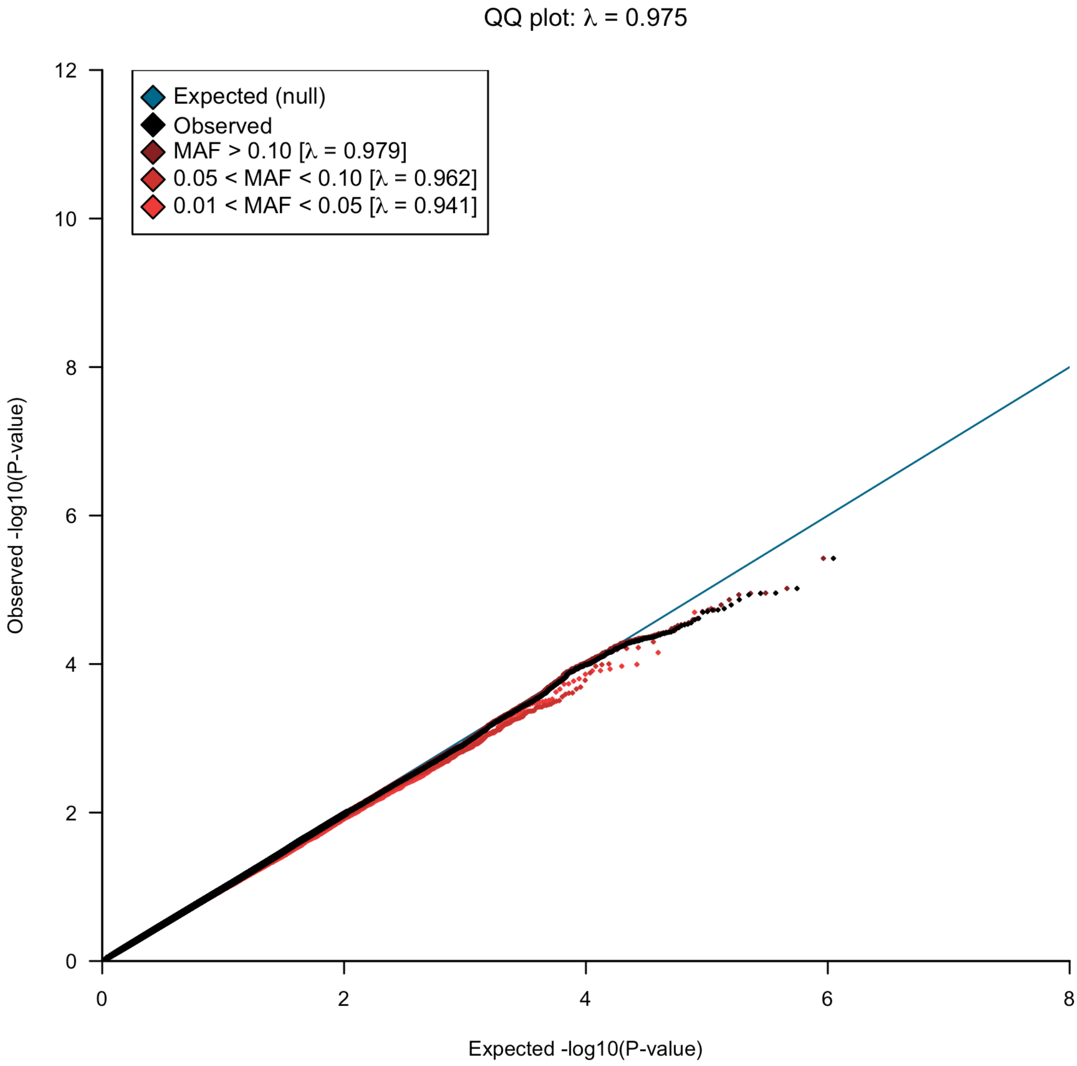


Supplementary Fig. S2. Quantile-quantile plot of the observed -$\log_{10}$[p-values] from Netherlands Twin Register (NTR) GWAS for age at onset of walking against the expected -$\log_{10}$[two-sided p-value] (blue line). Results are presented by allele frequency, brown dots representing signals for the SNPs with minor allele frequency (MAF) > 10%, red for SNPs with MAF between 5 and 10%, orange circles for SNPs with MAF between 1 and 5% and black circles for all SNPs included in the GWAS (MAF > 1%).

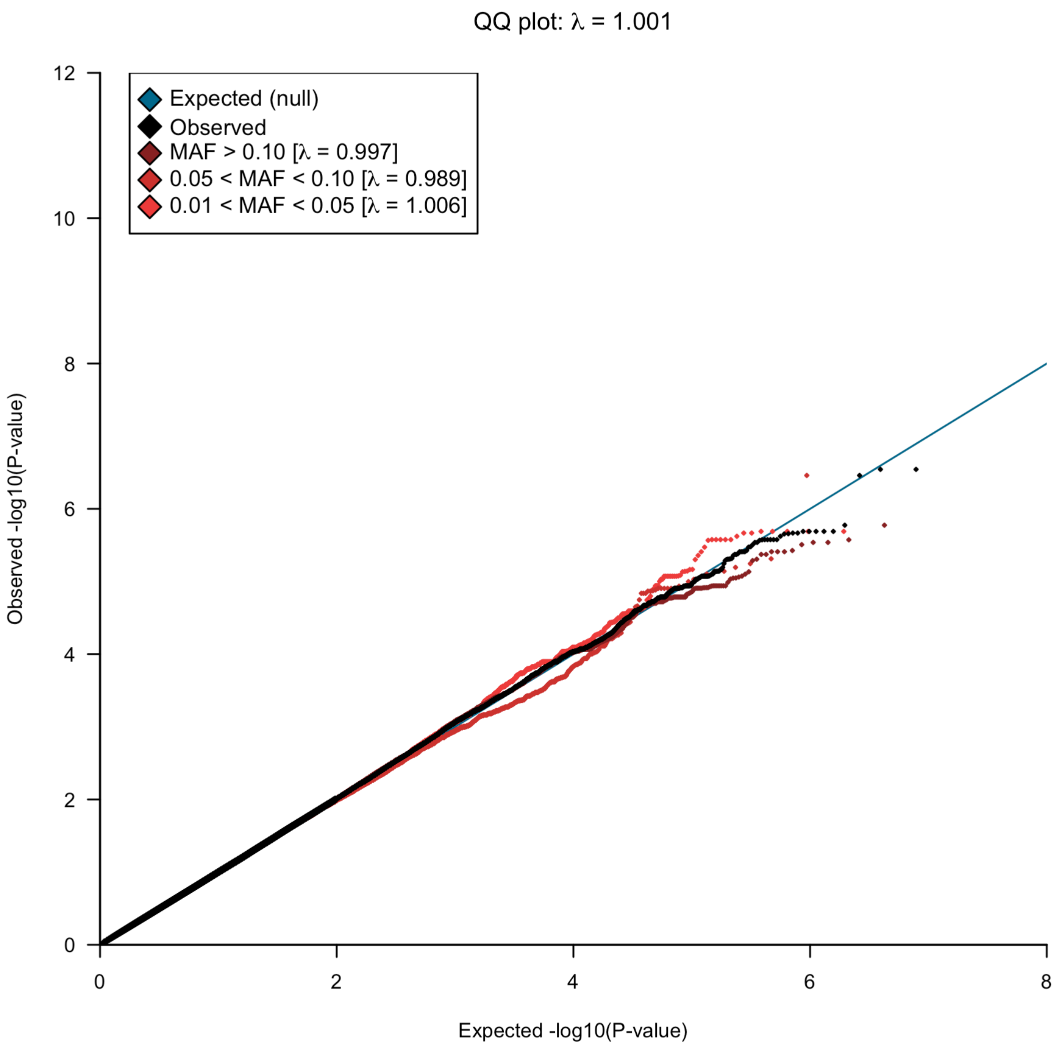


Supplementary Fig. S3. Quantile-quantile plot of the observed -$\log_{10}$[p-values] from Lifelines GWAS for age at onset of walking against the expected -$\log_{10}$[two-sided p-value] (blue line). Results are presented by allele frequency, brown dots representing signals for the SNPs with minor allele frequency (MAF) > 10%, red for SNPs with MAF between 5 and 10%, orange circles for SNPs with MAF between 1 and 5% and black circles for all SNPs included in the GWAS (MAF > 1%).

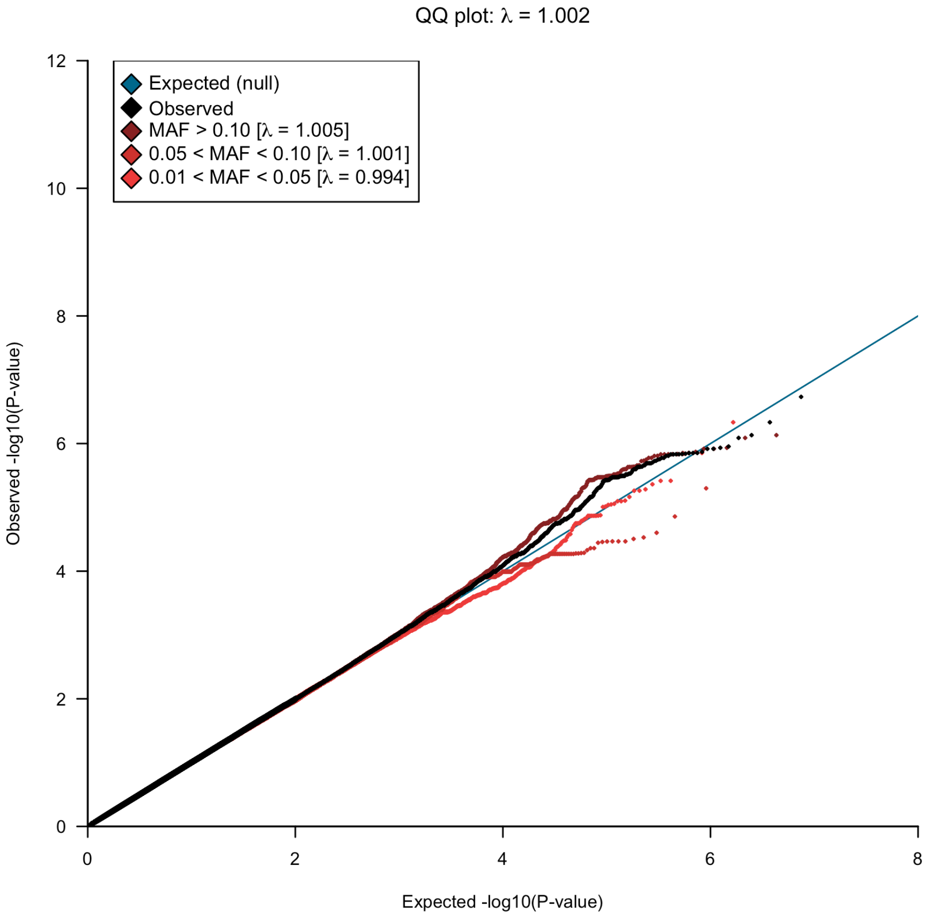


Supplementary Fig. S4. Quantile-quantile plot of the observed -$\log_{10}$[p-values] from MRC National Study of Health and Development (NSHD) GWAS for age at onset of walking against the expected -$\log_{10}$[two-sided p-value] (blue line). Results are presented by allele frequency, brown dots representing signals for the SNPs with minor allele frequency (MAF) > 10%, red for SNPs with MAF between 5 and 10%, orange circles for SNPs with MAF between 1 and 5% and black circles for all SNPs included in the GWAS (MAF > 1%).

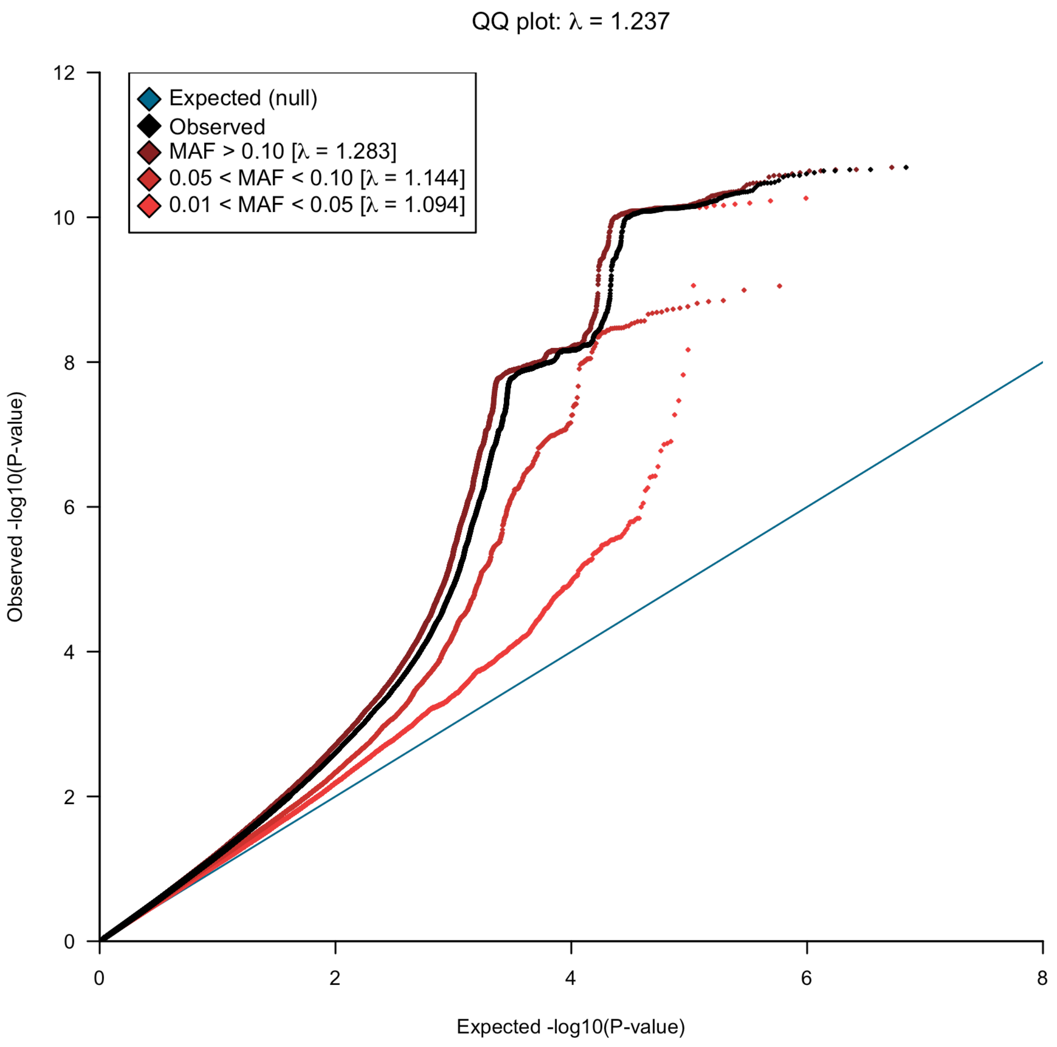


Supplementary Fig. S5. Quantile-quantile plot of the observed -$\log_{10}$[p-values] from GWAS meta-analysis for age at onset of walking (N=70,560) against the expected -$\log_{10}$[two-sided p-value] (blue line). Results are presented by allele frequency, brown dots representing signals for the SNPs with minor allele frequency (MAF) > 10%, red for SNPs with MAF between 5 and 10%, orange circles for SNPs with MAF between 1 and 5% and black circles for all SNPs included in the GWAS (MAF > 1%).

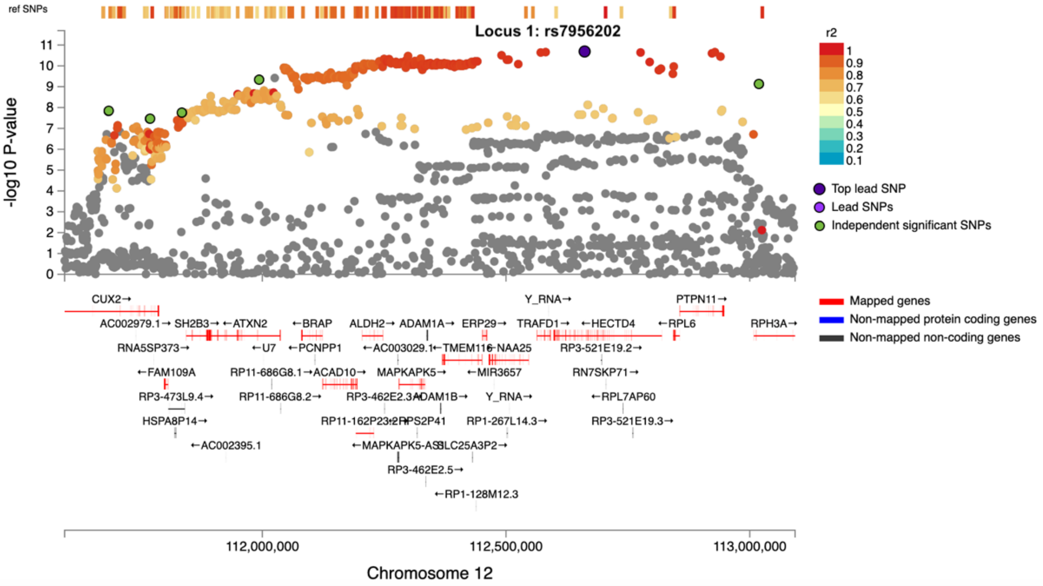
ref SNPs
Locus 1: rs7956202
-log10 P-value
r2
Top lead SNP
Lead SNPs
Independent significant SNPs
Mapped genes
Non-mapped protein coding genes
Non-mapped non-coding genes
CUX2→
Y_RNA→
PTPN11→
AC002979.1→
SH2B3→
←ATXN2
←BRAP
ALDH2→
ADAM1A→
ERP29→
TRAFD1→
←HECTD4
←RPL6
RPH3A→
RNA5SP373→
←U7
←PCNPP1
←AC003029.1
RP3-521E19.2→
←FAM109A
RP11-686G8.1→
ACAD10→
MAPKAPK5→
←MIR3657
RN7SKP71→
RP3-473L9.4→
RP11-686G8.2→
Y_RNA→
←RPL7AP60
HSPA8P14→
RP1-267L14.3→
RP3-521E19.3→
←AC002395.1
RP3-462E2.5→
←RP1-128M12.3
112,000,000
112,500,000
113,000,000
Chromosome 12

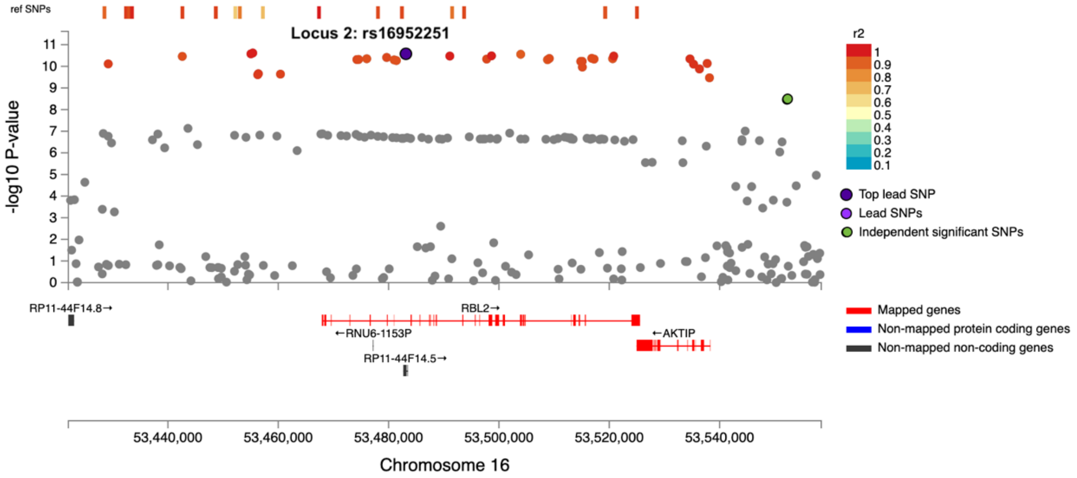
ref SNPs
Locus 2: rs16952251
-log10 P-value
r2
Top lead SNP
Lead SNPs
Independent significant SNPs
Mapped genes
Non-mapped protein coding genes
Non-mapped non-coding genes
RP11-44F14.8→
RBL2→
←RNU6-1153P
←AKTIP
RP11-44F14.5→
53,440,000
53,460,000
53,480,000
53,500,000
53,520,000
53,540,000
Chromosome 16

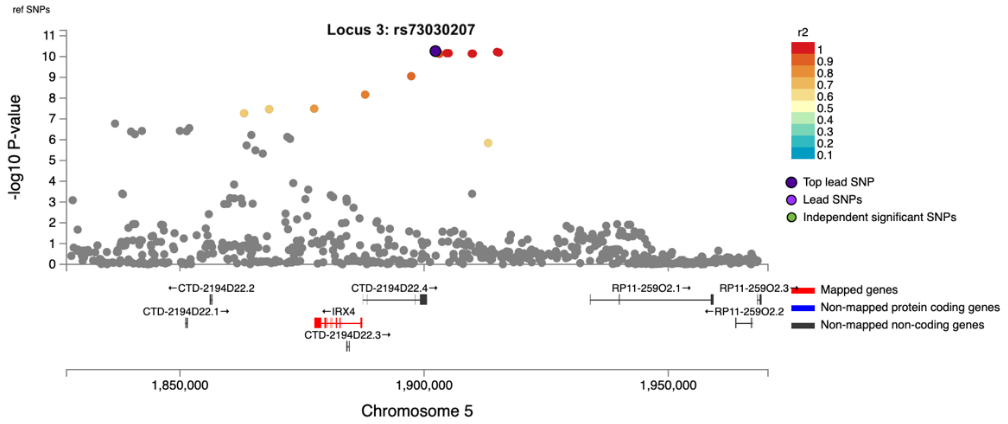
ref SNPs
Locus 3: rs73030207
-log10 P-value
r2
1
0.9
0.8
0.7
0.6
0.5
0.4
0.3
0.2
0.1
Top lead SNP
Lead SNPs
Independent significant SNPs
←CTD-2194D22.2
CTD-2194D22.4→
RP11-259O2.1→
RP11-259O2.3→
CTD-2194D22.1→
←IRX4
←RP11-259O2.2
CTD-2194D22.3→
Mapped genes
Non-mapped protein coding genes
Non-mapped non-coding genes
1,850,000
1,900,000
1,950,000
Chromosome 5

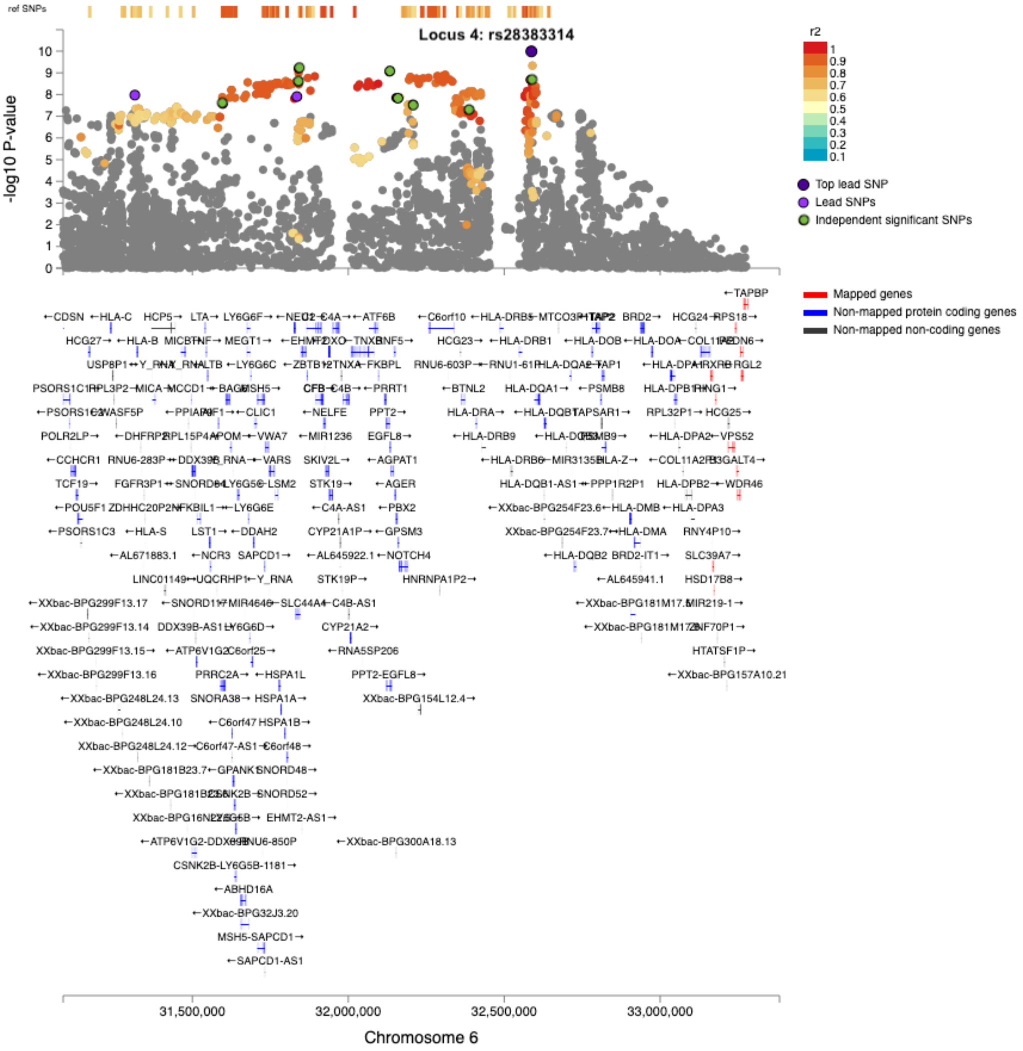
ref SNPs
Locus 4: rs28383314
-log10 P-value
r2
1
0.9
0.8
0.7
0.6
0.5
0.4
0.3
0.2
0.1
Top lead SNP
Lead SNPs
Independent significant SNPs
Mapped genes
Non-mapped protein coding genes
Non-mapped non-coding genes
31,500,000
32,000,000
32,500,000
33,000,000
Chromosome 6

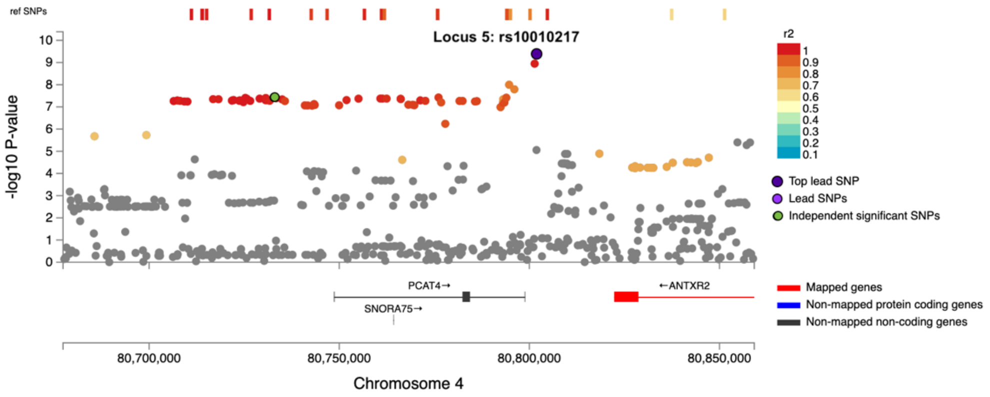
ref SNPs
Locus 5: rs10010217
-log10 P-value
r2
1
0.9
0.8
0.7
0.6
0.5
0.4
0.3
0.2
0.1
Top lead SNP
Lead SNPs
Independent significant SNPs
PCAT4→
SNORA75→
←ANTXR2
Mapped genes
Non-mapped protein coding genes
Non-mapped non-coding genes
80,700,000
80,750,000
80,800,000
80,850,000
Chromosome 4

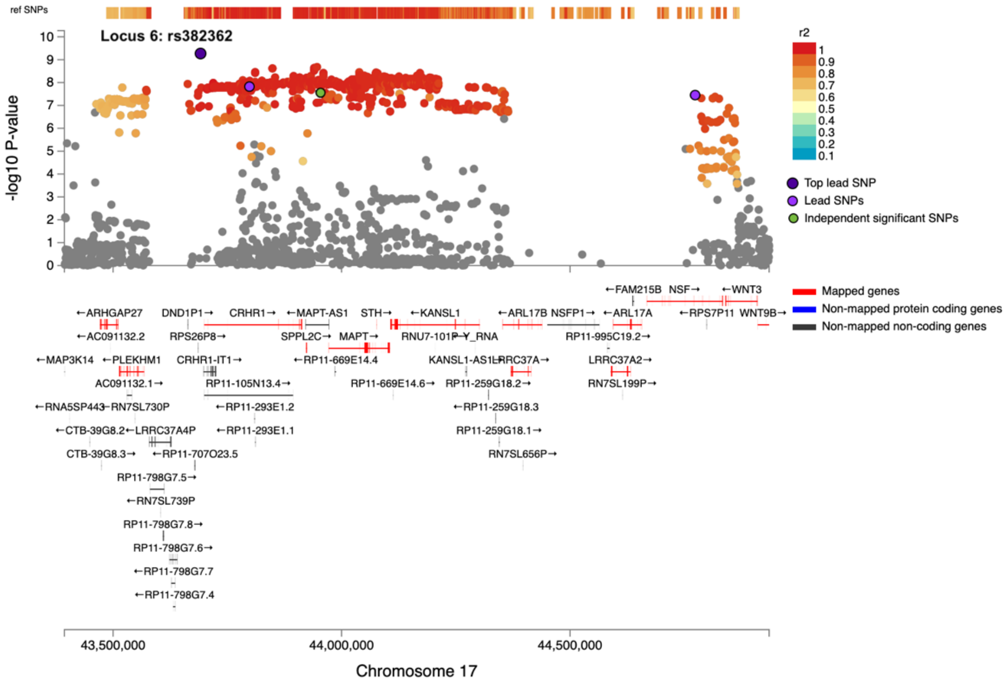
ref SNPs
Locus 6: rs382362
-log10 P-value
r2
Top lead SNP
Lead SNPs
Independent significant SNPs
Mapped genes
Non-mapped protein coding genes
Non-mapped non-coding genes
←FAM215B NSF→ ←WNT3
←ARHGAP27 DND1P1→ CRHR1→ ←MAPT-AS1 STH→ ←KANSL1 ←ARL17B NSFP1→ ←ARL17A ←RPS7P11 WNT9B→
←AC091132.2 RPS26P8→ SPPL2C→ MAPT→ RNU7-101P Y_RNA RP11-995C19.2→
←MAP3K14 ←PLEKHM1 CRHR1-IT1→ ←RP11-669E14.4 KANSL1-AS1 LRRC37A→ LRRC37A2→
AC091132.1→ RP11-105N13.4→ RP11-669E14.6→ RP11-259G18.2→ RN7SL199P→
←RNA5SP443 RN7SL730P ←RP11-293E1.2 ←RP11-259G18.3
←CTB-39G8.2 ←LRRC37A4P ←RP11-293E1.1 RP11-259G18.1→
CTB-39G8.3→ ←RP11-707O23.5 RN7SL656P→
RP11-798G7.5→
←RN7SL739P
RP11-798G7.8→
RP11-798G7.6→
←RP11-798G7.7
←RP11-798G7.4
43,500,000
44,000,000
44,500,000
Chromosome 17

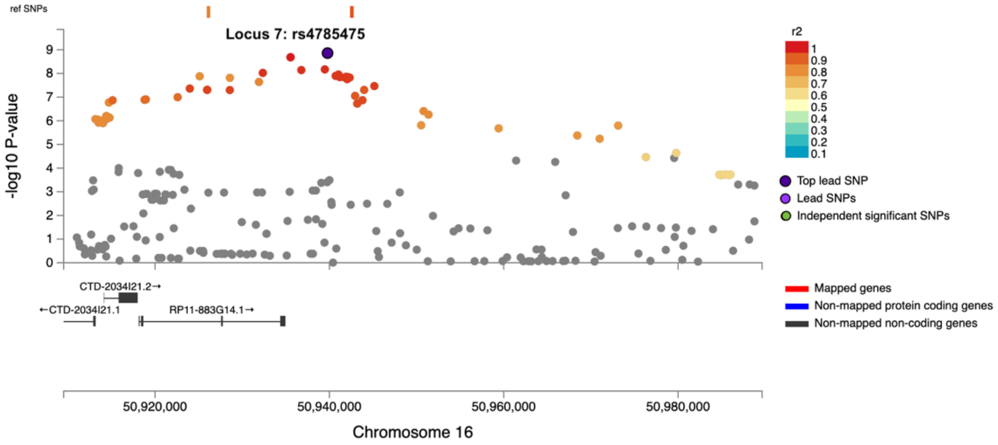
ref SNPs
Locus 7: rs4785475
-log10 P-value
r2
Top lead SNP
Lead SNPs
Independent significant SNPs
CTD-2034I21.2→
←CTD-2034I21.1
RP11-883G14.1→
Mapped genes
Non-mapped protein coding genes
Non-mapped non-coding genes
50,920,000
50,940,000
50,960,000
50,980,000
Chromosome 16

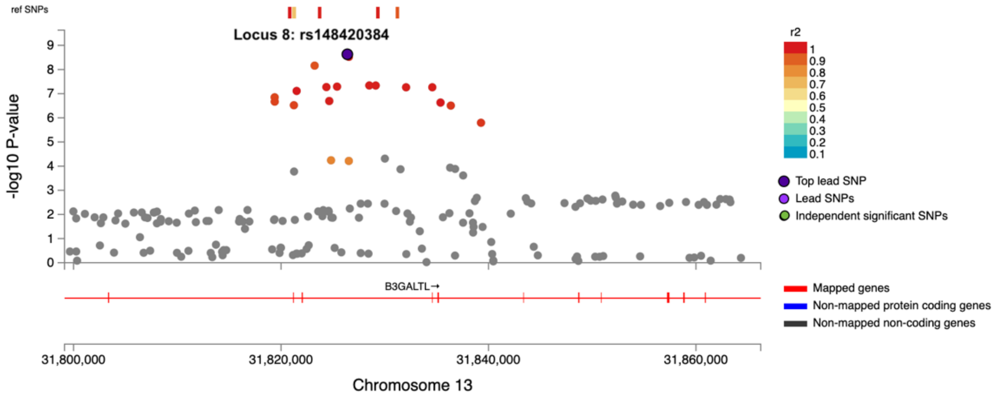
ref SNPs
Locus 8: rs148420384
-log10 P-value
r2
1
0.9
0.8
0.7
0.6
0.5
0.4
0.3
0.2
0.1
Top lead SNP
Lead SNPs
Independent significant SNPs
B3GALTL→
Mapped genes
Non-mapped protein coding genes
Non-mapped non-coding genes
31,800,000
31,820,000
31,840,000
31,860,000
Chromosome 13

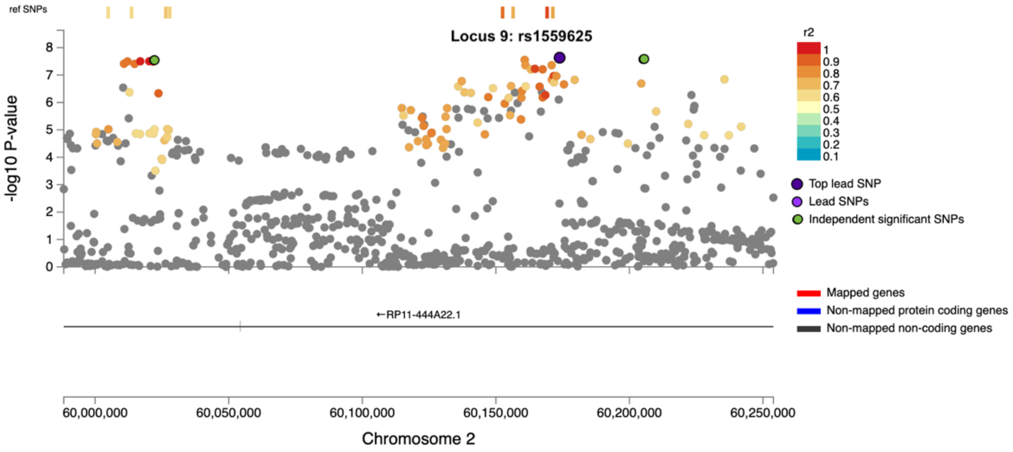
ref SNPs
Locus 9: rs1559625
-log10 P-value
r2
1
0.9
0.8
0.7
0.6
0.5
0.4
0.3
0.2
0.1
Top lead SNP
Lead SNPs
Independent significant SNPs
←RP11-444A22.1
Mapped genes
Non-mapped protein coding genes
Non-mapped non-coding genes
60,000,000
60,050,000
60,100,000
60,150,000
60,200,000
60,250,000
Chromosome 2

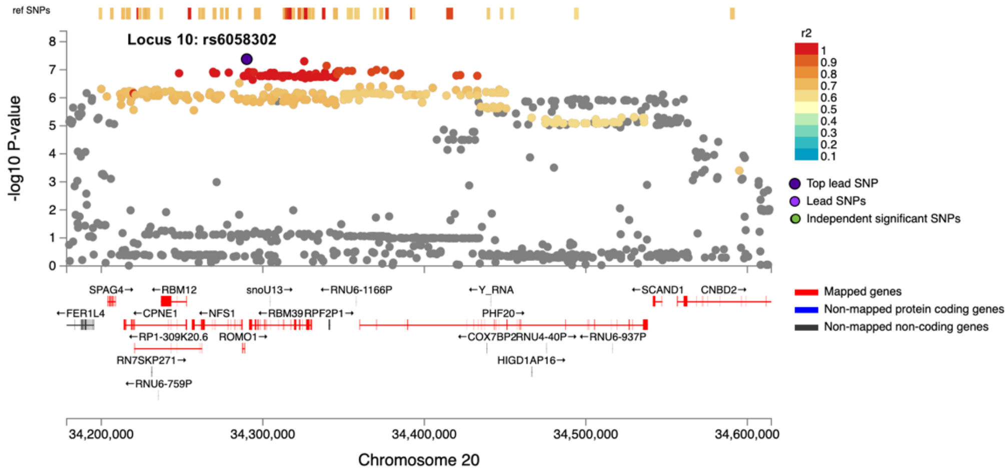
ref SNPs
Locus 10: rs6058302
-log10 P-value
r2
1
0.9
0.8
0.7
0.6
0.5
0.4
0.3
0.2
0.1
Top lead SNP
Lead SNPs
Independent significant SNPs
SPAG4→
←RBM12
snoU13→
←RNU6-1166P
←Y_RNA
←SCAND1
CNBD2→
←FER1L4
←CPNE1
←NFS1
←RBM39
RPF2P1→
PHF20→
←RP1-309K20.6
ROMO1→
←COX7BP2
RNU4-40P→
←RNU6-937P
RN7SKP271→
HIGD1AP16→
←RNU6-759P
Mapped genes
Non-mapped protein coding genes
Non-mapped non-coding genes
34,200,000
34,300,000
34,400,000
34,500,000
34,600,000
Chromosome 20

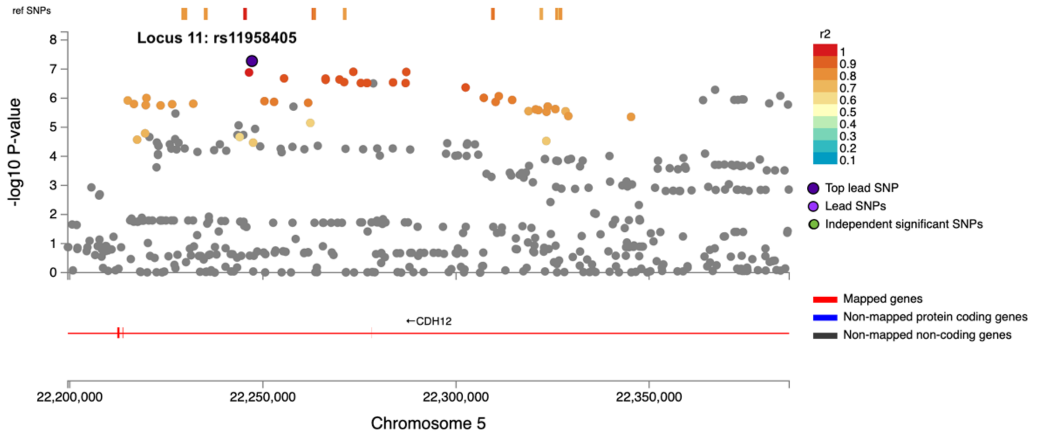


Supplementary Fig. S6. Regional plots for the top SNPs in the eleven genomic loci associated with age at onset of walking, realised with FUMA[27]. Association p-values on the $-\log_{10}$ scale are shown on the y axis, and the chromosomal position is indicated along the x axis. The top lead SNP within the locus is indexed by a blue dot and annotated with the genomic locus number and SNP ID. Other independent (LD $r^2$<0.6) significant SNPs are indexed in green. SNPs in LD with the lead variant are color-coded based on LD $r^2$ estimated on the 1000 Genomes[15] European population. SNPs that are not in LD with any of the significant independent lead SNPs in the region are colored grey. At the bottom, genes mapped by positional mapping are presented in red.

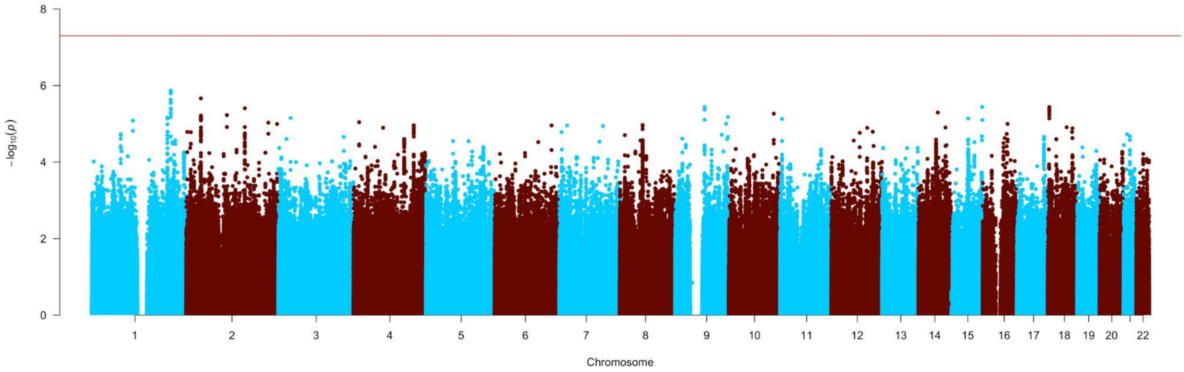


Supplementary Fig. S7. Heterogeneity Manhattan plot. The y-axis represents the -$\log_{10}$[two-sided p-value] of an omnibus test for heterogeneity across cohorts ($I^2$ statistics) for the age at onset of walking meta-GWAS. The x-axis represents genomic position (chromosomes 1-22). The horizontal red line indicates the p-value threshold for genome-wide statistical significance ($p = 5 \times 10^{-8}$).

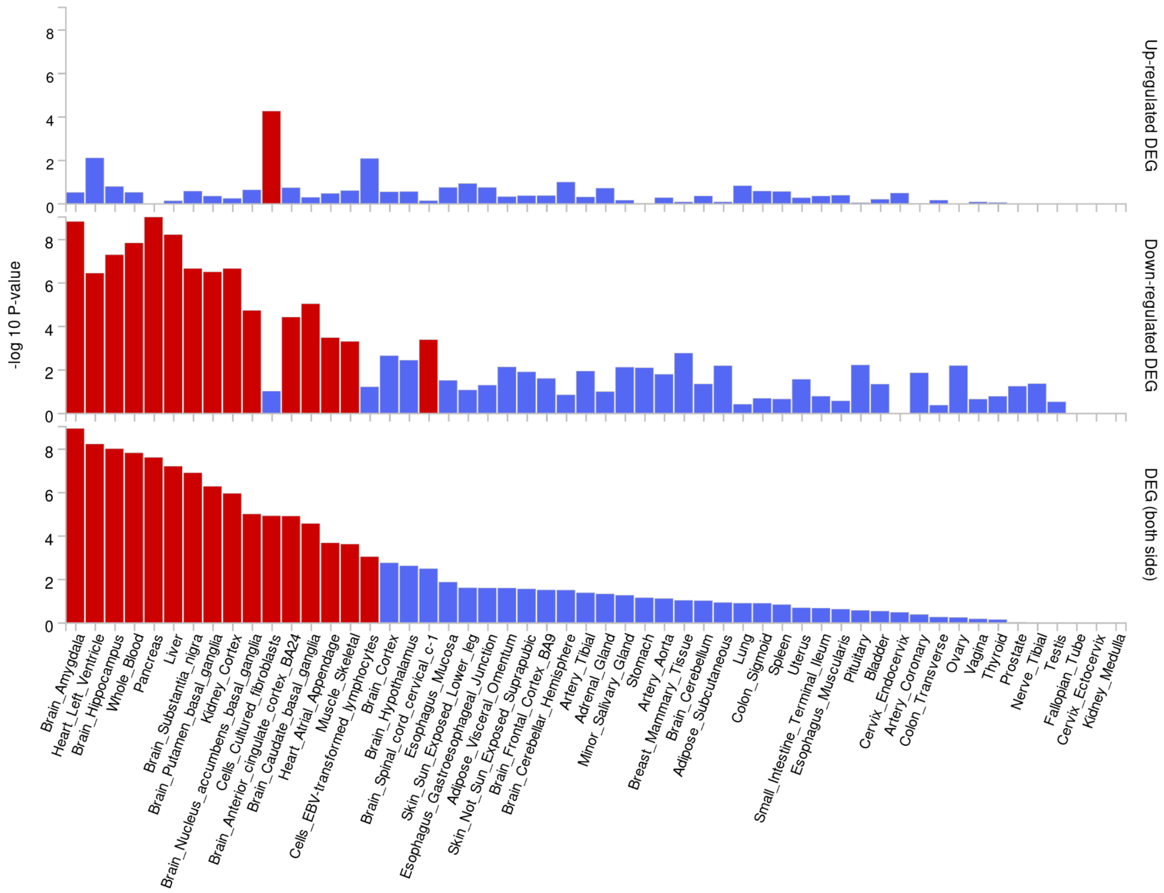


Supplementary Fig. S8. Enrichment of up-regulated and down-regulated differentially expressed genes (DEGs) performed for 54 GTEx[29] v8 tissue datasets available in FUMA[27] (on the x axis). The y axis represents the -$\log_{10}$[one-sided p-value] of the probability hypergeometric test performed in FUMA (GENE2FUNC). Red bars indicate the tissue for which the DEGs are enriched at a Bonferroni corrected p-value < 0.05.

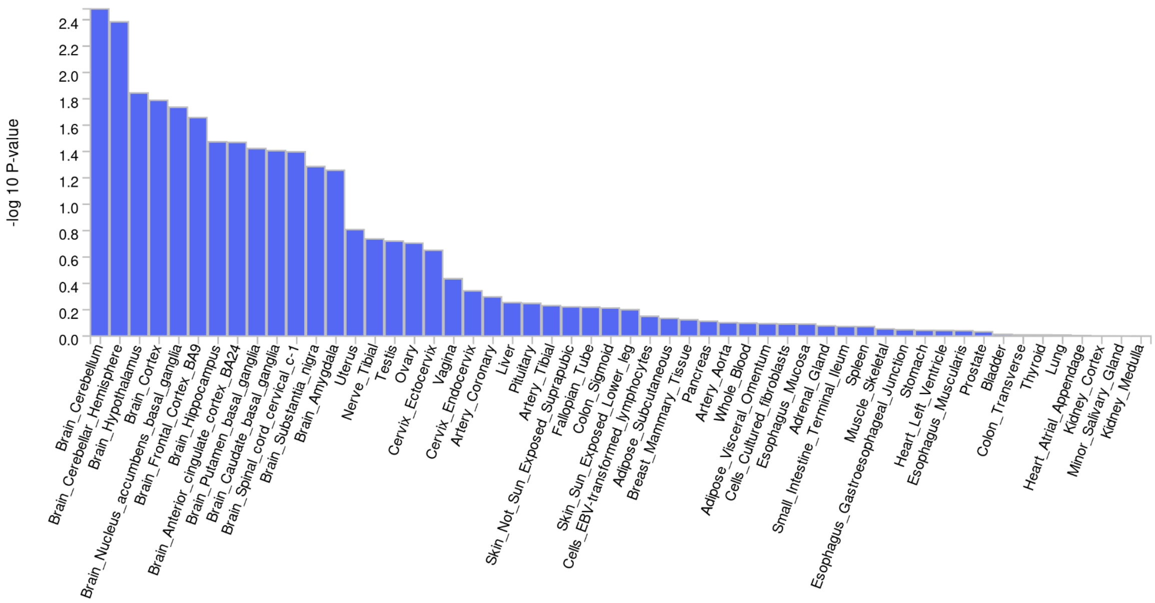


Supplementary Fig. S9. Results of the MAGMA[30] gene property analysis performed for 53 GTEx[29] v8 tissue types (on the x axis). The y axis represents the $-\log_{10}$[one-sided p-value] testing the positive relationship between tissue specificity and genetic association of genes with age at onset of walking.

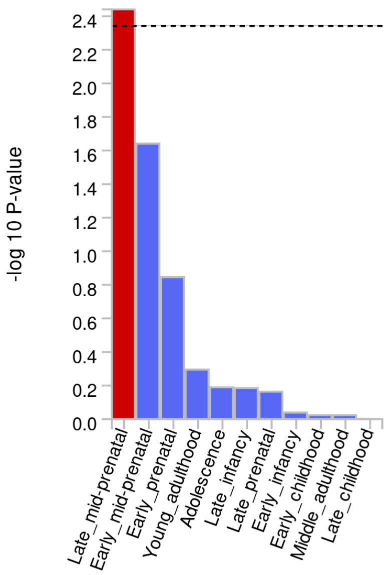


Supplementary Fig. S10. Results of the MAGMA[30] gene property analysis performed for the BrainSpan developmental stages dataset[28] available in FUMA[27] (on the x axis). The y axis represents the -$\log_{10}$[one-sided p-value] testing the positive relationship between expression in the BrainSpan tissues and genetic association of genes with age at onset of walking. The dashed line represents the Bonferroni-corrected α level, and the red bars indicate that the beta-value of the regression between gene expression in the BrainSpan tissues and genetic association with age at onset of walking is significantly higher than 0.

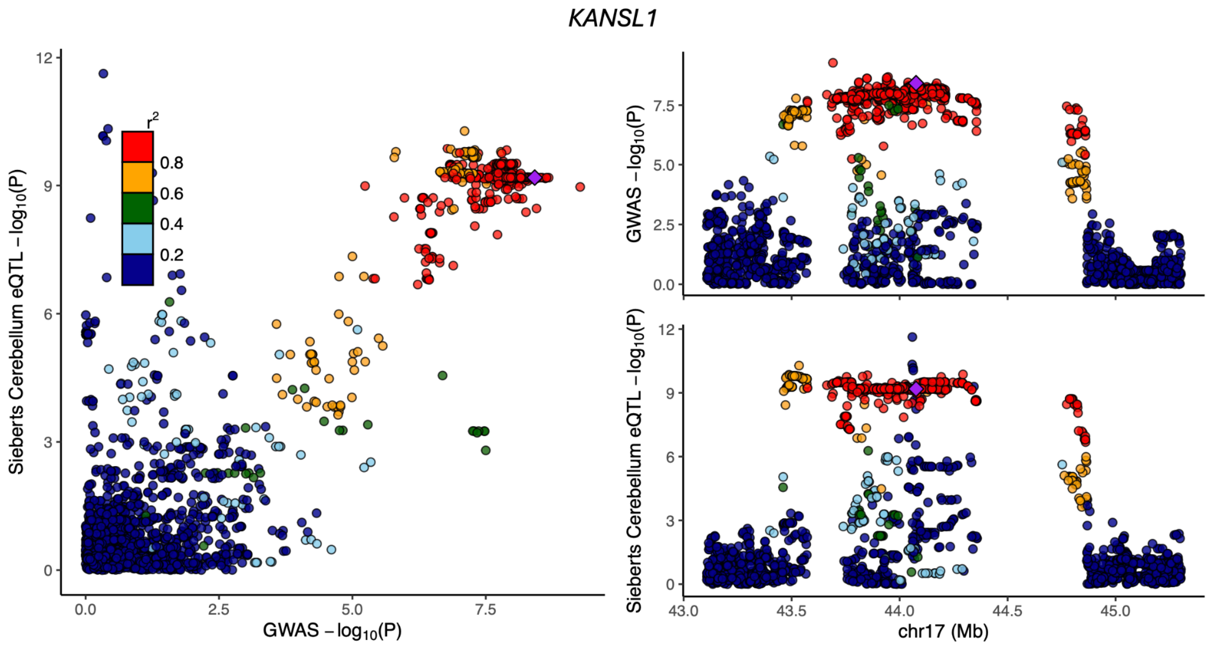


Supplementary Fig. S11. GWAS evidence for association with age at onset of walking (-$\log_{10}$[p-value], *x*-axis) plotted against the statistical evidence of being an expression Quantitative Trait Locus (eQTL) for gene *KANSL1* within the genomic locus 6 in human adult cerebellum[31] (-$\log_{10}$[p-value]), *y*-axis) for each SNP (points) within a 1Mb window around the gene. Points are colored by LD correlation with the lead SNP (purple diamonds), estimated in individuals with European ancestry in 1000 Genomes[15,32].

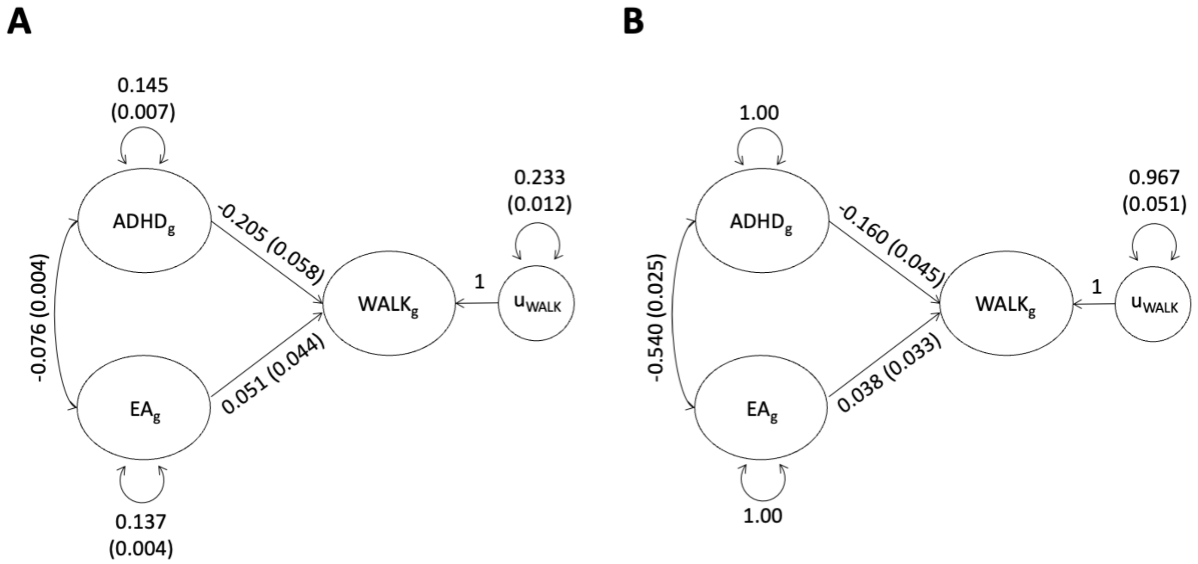


Supplementary Fig. S12. Unstandardized (A) and standardized (B) results from Genomic SEM[33] indicating the genetic covariance and genetic correlation estimates, respectively, and associated sampling covariance matrices, from a multivariate LD score regression of the age at learning to walk (WALK) on ADHD[8] and educational attainment (EA)[34].

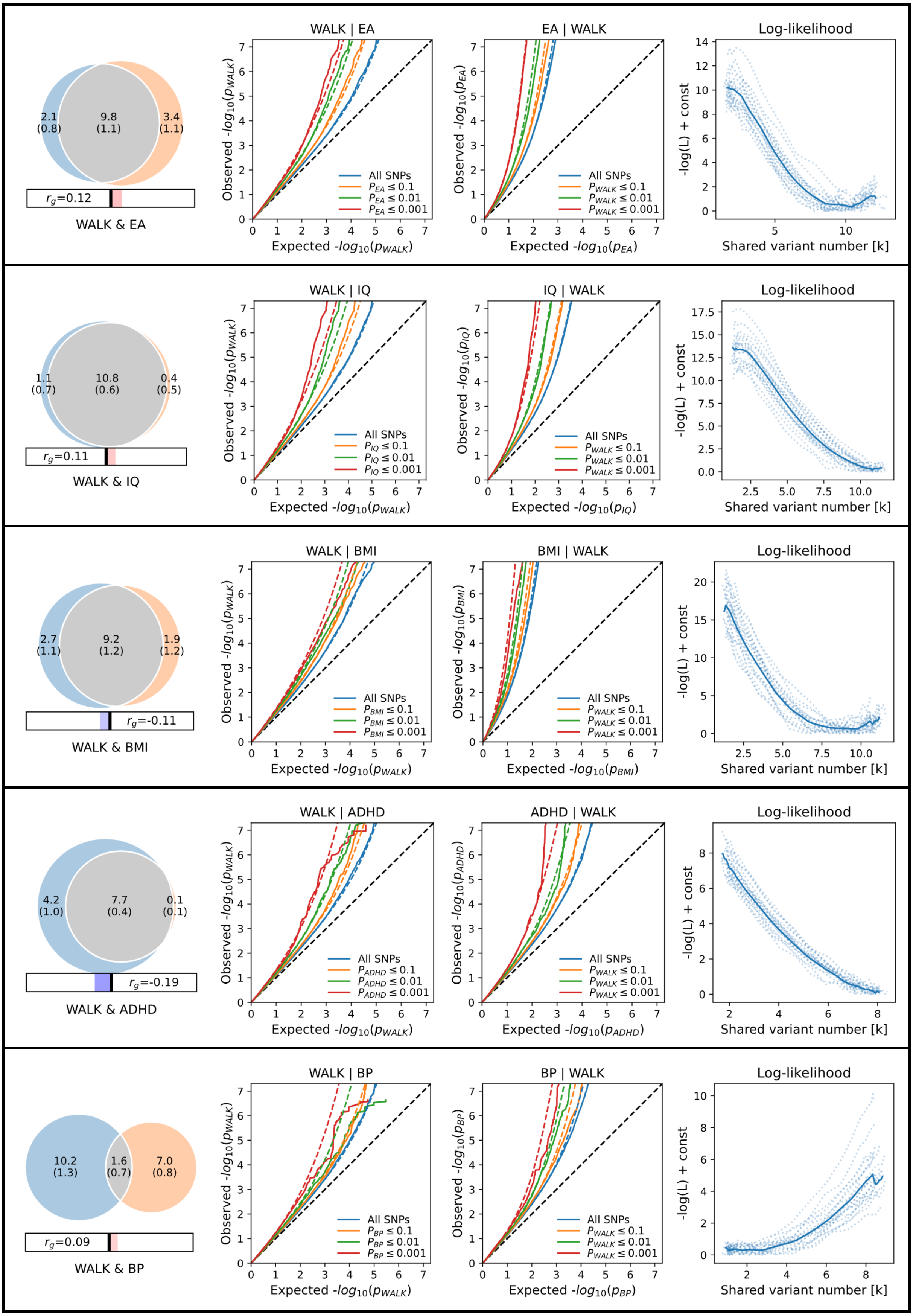

WALK & EA
$r_g$=0.12
WALK | EA
EA | WALK
Log-likelihood
WALK & IQ
$r_g$=0.11
WALK | IQ
IQ | WALK
WALK & BMI
$r_g$=-0.11
WALK | BMI
BMI | WALK
WALK & ADHD
$r_g$=-0.19
WALK | ADHD
ADHD | WALK
WALK & BP
$r_g$=0.09
WALK | BP
BP | WALK
All SNPs
Expected -$log_{10}(p_{WALK})$
Observed -$log_{10}(p_{WALK})$
-log(L) + const
Shared variant number [k]

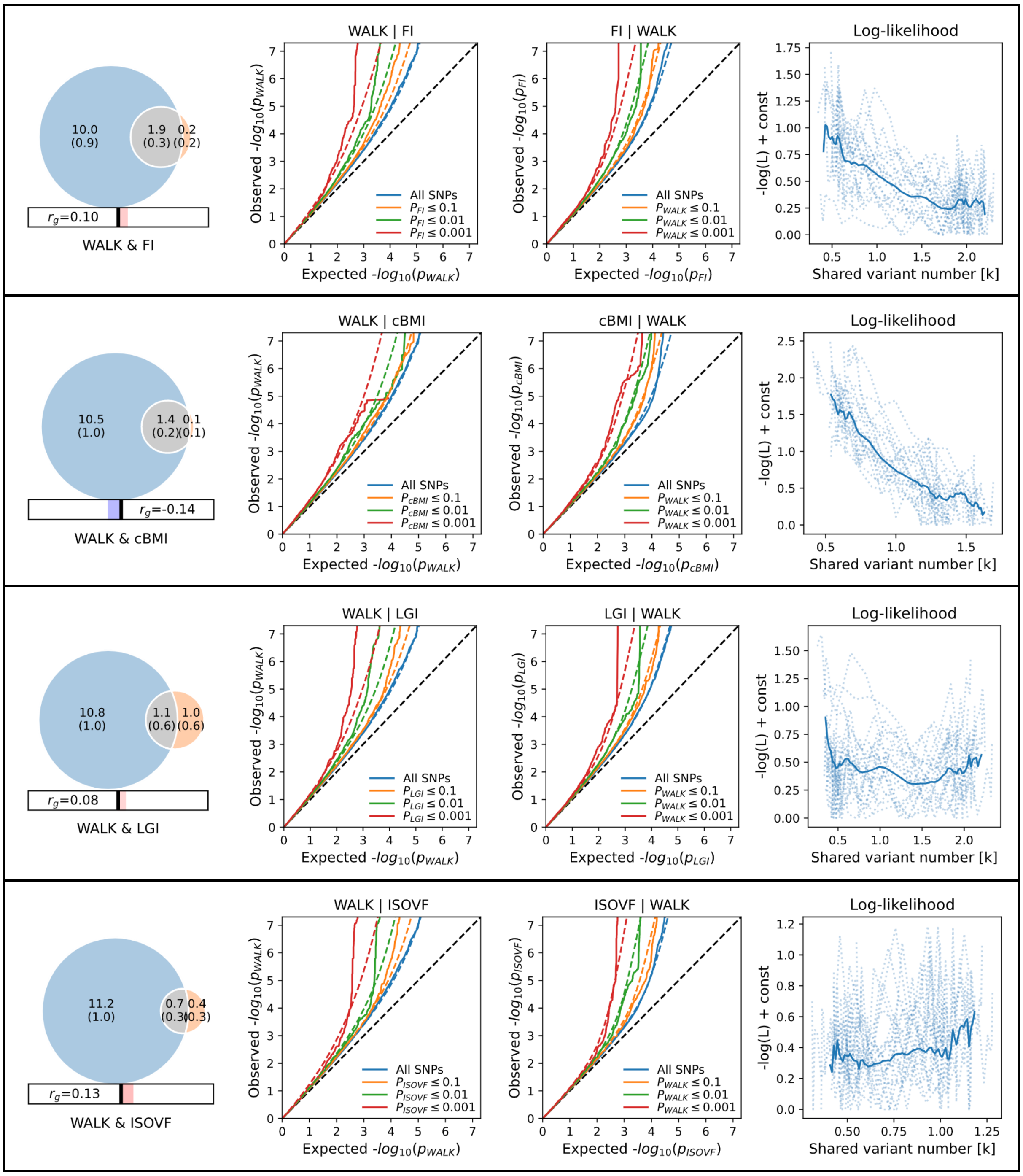


Supplementary Fig. S13. MiXeR[7] bivariate model figures including negative log-likelihood plots. WALK = age at onset of walking; IQ = Intelligence; BMI = Body Mass Index; ADHD = Attention Deficit / Hyperactivity Disorder; BP = Bipolar Disorder; FI = Folding Index; cBMI = childhood BMI; LGI = Local Gyrification Index; ISOVF = Isotropic Volume Fractionation.
